# Serum proteins predict incident inflammatory bowel disease 14 years before diagnosis: a machine-learning and Cox modelling in the UK Biobank

**DOI:** 10.64898/2026.10.13.26365425

**Authors:** Tobias Erich Niebuhr, Joshua Fieggen, Bradley Max Segal, Anuj Patel, David Clifton, Lei Clifton

## Abstract

Using an analysis pipeline that combines machine learning-based feature selection with classical statistical modelling, we aimed to predict incident inflammatory bowel disease (IBD) and to identify candidate proteins associated with disease development. We used UK Biobank data in an observational cohort study of 52,331 participants with proteomics data and without a history of IBD. The cohort was split into an 80/20 train/test dataset, and three separate extreme gradient boosting models were trained on the training dataset. Using Shapley values, we ranked the predictive importance of 2,912 proteins for incident IBD, Crohn’s disease (CD), and ulcerative colitis (UC). The top 20 proteins per outcome served as features in multivariate Cox models predicting 350 incident cases over 14 years. All three proteomics Cox models showed moderate discrimination in the test dataset, with the UC model performing the best (C-index: 0.68), followed by IBD (C-index: 0.65), and CD (C-index: 0.60). All proteomics models outperformed clinical models. Smoking remained the only significant clinical predictor in the combined proteomics + clinical model. 48 unique proteins were identified across the three models, with 16 reaching statistical significance (p< 0.05). Machine learning-based feature selection with statistical survival modelling identifies plasma proteins that predict incident IBD and highlight biologically plausible, potentially targetable pathways in the preclinical phase of IBD.

## Introduction

Inflammatory bowel disease (IBD) is a chronic immune-mediated condition of the gastrointestinal tract, characterised by relapsing and remitting intestinal inflammation that causes severe morbidity^1–4^. IBD affects an estimated 4.9 million people globally, and consists of two major subtypes, Crohn’s disease (CD) and ulcerative colitis (UC), with UC being about twice as prevalent as CD^5^. Although CD and UC differ in their clinical presentation and disease course^2^, recent evidence suggests that they lie on a spectrum of disease^6,7^. There is no known cure for IBD and, despite treatment with immunosuppressive medications, many patients will live with substantial symptom burden, frequent hospitalisations, and even surgery^8,9^.

The incidence of IBD is rising disproportionately in developing countries^2,5^ and, through earlier diagnosis, an ageing population, and less IBD-related mortality^1^, IBD will soon become a considerable global health burden, but minimal progress has been made in prediction and prevention as the underlying aetiology is still poorly understood.

According to their recent workshop on the prediction and prevention of IBD^1^, the European Crohn’s and Colitis Organisation (ECCO) found evidence that proteins are associated with dysregulated immune pathways during the preclinical stage, years before clinically diagnosable symptoms^1,4,10,11^. ECCO identified accurate predictive models in the preclinical phase as a top research priority, as studying those proteins could allow earlier detection of those likely to develop IBD, a deeper understanding of the underlying inflammatory processes, and could lead to drug discovery tailored to preventing progression from preclinical to clinical IBD^1^.

Proteomics has been used to predict IBD and identify associated proteins. In the Nurses’ Health Study cohort, Lochhead et al.^11^ found that higher C-reactive protein and interleukin-6 levels are associated with an increased risk of developing IBD, and Torres et al. found 51 proteins that are predictive of CD in the PREDICTS cohort, with a 76% accuracy in predicting CD diagnosis in 5 years, and 87% in 1 year^10^.

In the Canadian GEM cohort, Leibovitzh et al.^12^ found 25 serum proteins that are associated with developing CD, three of which were also associated with developing UC in the Swedish National Registry^13^, suggesting a shared inflammatory pathway. Kalla et al.^14^ similarly found that most proteins predictive of CD and UC are shared between the two, indicating a shared pathophysiology.

Most of these studies were limited by a narrow proteomic coverage or small sample sizes, and relied on classical statistical methods. As proteomic assays develop to measure thousands of proteins simultaneously, classical statistical methods increasingly struggle to handle this level of dimensionality robustly. Machine learning (ML) techniques often outperform them in such settings, while also being able to identify complex non-linear relationships between proteins and the outcome that classical models might miss^15,16^.

In 2025, two studies used ML and statistical methods in the UK Biobank (UKB) to predict IBD up to 16 years before diagnosis, using explainability techniques to explain which proteins are most predictive of developing IBD^17^, as well as the subtypes CD^17,18^ and UC^17^.

Both studies used univariate Cox models for feature selection, a filter method that is often used to reduce dimensionality in large datasets. While being simple and scalable, this method generally ignores interactions between features and is prone to false positive discoveries through multiple testing, as it selects only features that are significantly associated with the outcome^19^.

Following this, both studies trained extreme gradient boosting (XGBoost) models on the reduced dataset to predict IBD^17^, CD^17,18^ and UC^17^. They ranked proteins in order of their Shapley Additive exPlanations (SHAP) values and selected the top 10 proteins for each model, which were then used as features in a simplified tree-based model to provide maximum performance with a small amount of features.

Feng et al. achieving a good discriminatory ability in their 9-protein model at 16 years for developing CD (AUROC = 0.72)^18^. In comparison, Zhang et al.’s pipeline for CD did not select any of the same proteins and had a poorer performance (AUROC = 0.62)^17^. Zhang et al. developed separate IBD and UC models, in which the IBD model showed the best performance for all-time IBD incidence (AUROC = 0.67). Notably, both Feng et al. and Zhang et al. also constructed a clinical-only model and a combined model (proteomic plus clinical features) for each disease which performed worse across all disease groups.

By relying on filter methods, however, they fail to capture complex interactions between proteins, and do not leverage the main advantage of XGBoost’s embedded feature selection, namely selecting from thousands of features during model training while identifying non-linear relationships. Their SHAP values also quantify contribution to the model’s predictions rather than the magnitude and direction of effect, so they cannot be used to draw conclusions about statistical association^16,20^.

In this study, we analysed 2,912 plasma proteins measured in 52,331 participants of the UK Biobank (UKB), a prospective population-based cohort followed for up to 16 years, to identify proteins predictive of incident IBD and its subtypes. We trained XGBoost models on the full proteomic space and used their embedded feature importance through SHAP values to select the most predictive proteins. Unlike conventional linear approaches, this method can capture non-linear relationships, which may better reflect inflammatory processes in IBD. Using this approach, we derived candidate protein panels for IBD, as well as its subtypes CD, and UC. To keep the results statistically interpretable, we estimated the association of each selected protein with time to diagnosis using Cox proportional hazards models. We then compared the selected proteins with the existing literature to interpret the biological pathways potentially involved in disease development. Finally, we highlighted proteins with no prior reported link to IBD as priorities for future investigation. An overview of the analysis pipeline is shown in Figure 1.

**Figure 1.**
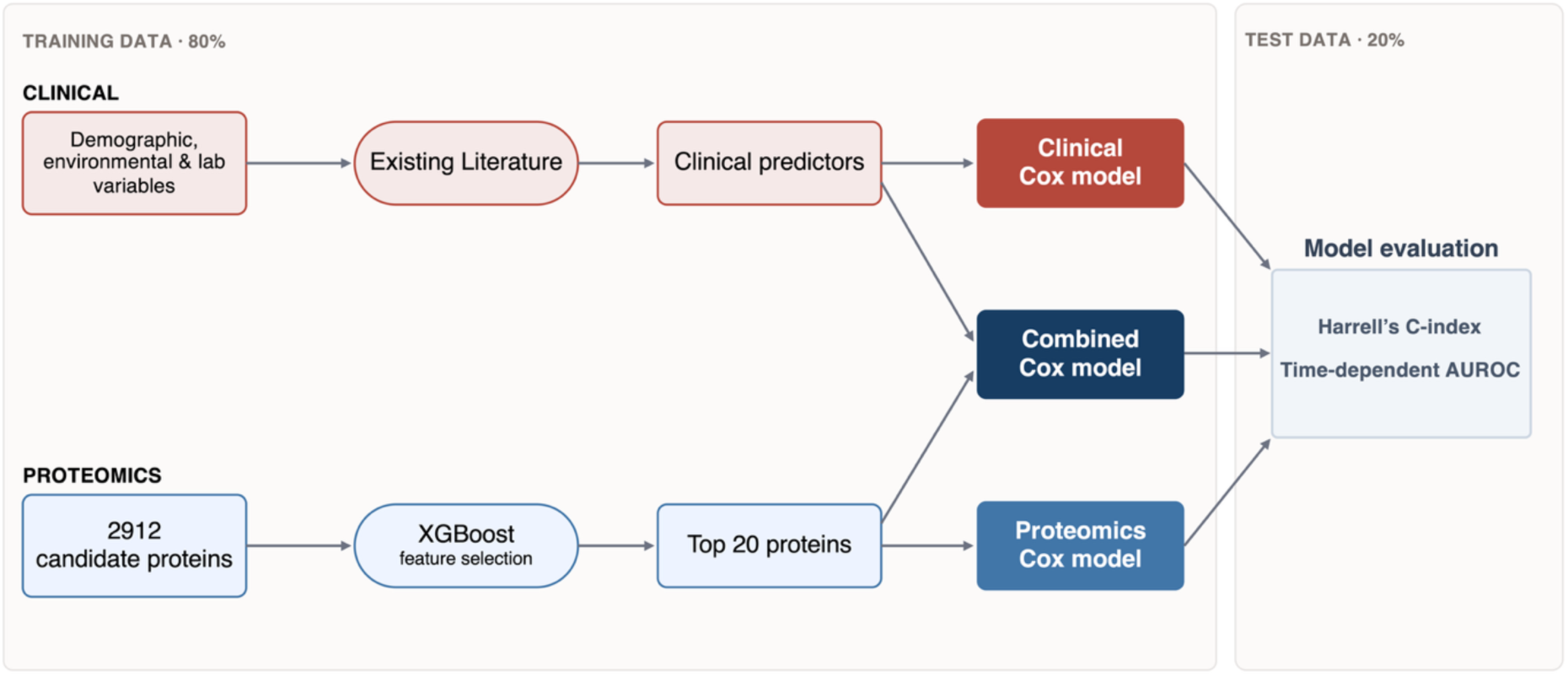
Analysis pipeline. XGBoost was used to select the top 20 protein predictors for developing IBD, which were used as covariates in Cox modelling. The analysis pipeline was repeated for the two subtypes, CD and UC. Three Cox models were constructed using clinical predictors, protein predictors, and a combination of the clinical and protein predictors. Model performance was assessed using Harrell’s C-index and time-dependent AUC. Harrell’s C-index is a time-independent performance metric measuring the models ability to assign a higher risk to cases than non-cases, and time-dependent AUC is a measure of a model’s predictive ability at defined time-points, assessing the performance trend over time. **XGBoost**: eXtreme Gradient Boosting machine, **AUROC**: Area under the Receiver Operator Curve

## Results

### Patient characteristics

A total of 501,932 participants were enrolled in the UKB cohort study. 5,827 participants with prevalent IBD were excluded (1,876 prevalent CD and 3,951 prevalent UC cases), and 443,774 participants were excluded due to missing proteomics data, leaving a study population of 52,331 participants with proteomics data available and no prior history of IBD (Figure 2).

**Figure 2.**
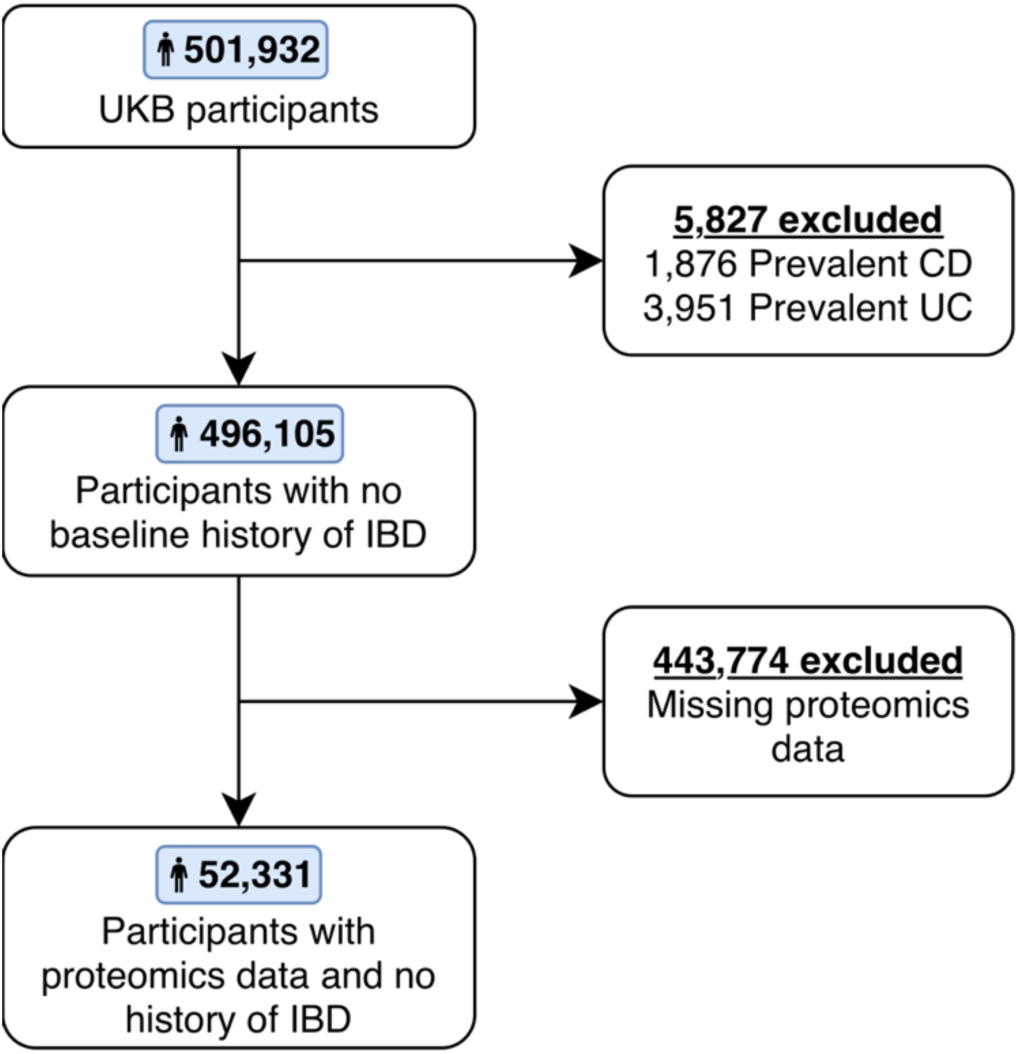
Selection of study participants.

Incident IBD was diagnosed in 350 participants (incidence rate: 0.66%) over a median follow-up time of 13.6 years (minimum: 0.01 years, maximum: 17.04 years). Of these, 116 (33.1%) were diagnosed with CD and 234 (66.9%) with UC, with an incidence rate of 0.22% and 0.44% respectively. The median time to diagnosis was 6.8 years (range 0.01 to 15.0 years). Follow-up for non-cases until censoring ranged from 0.02 years to 17.0 years, with a median follow-up time of 14.0 years.

Study participants were predominantly of white ethnicity, which was even more pronounced in IBD cases (93.2% vs 95.7%). Participants who went on to develop IBD were more likely to smoke, and their C-reactive protein (CRP) and triglyceride levels were higher than those of controls. Further baseline characteristics are summarised in Table 1.

**Table 1.** Baseline Characteristics of the study population by incident IBD status. Mean (SD) and Median [interquartile ranges, IQR] are reported for continuous variables, and frequency (percentage) are reported for categorical variables. N is the number of non-missing values.

|  | Without incident IBD Controls (n = 51,981) | With Incident IBD Cases (n = 350) | N |
| --- | --- | --- | --- |
| **Demographic features** |  |  |  |
| Age (Years) | 56.8 (8.2),<br>58.0 [50.0 - 64.0] | 58.2 (7.7),<br>60.0 [53.0 - 64.0] | 52,331 |
| Sex |  |  | 52,331 |
| Male | 23 934 (46.0%) | 167 (47.7%) | 24,101 |
| Female | 28 047 (54.0%) | 183 (52.3%) | 28,230 |
| Ethnicity |  |  | 52,331 |
| White | 48 437 (93.2%) | 335 (95.7%) | 48,772 |
| Black | 971 (1.9%) | 4 (1.1%) | 975 |
| Chinese | 1208 (2.3%) | 4 (1.1%) | 1212 |
| Other | 1 108 (2.1%) | 7 (2.0%) | 1115 |
| Unknown | 257 (0.5%) | 0 (0.0%) | 257 |
| Body mass index (kg/m<sup>2</sup>) | 27.5 (4.8),<br>26.8 [24.2 - 29.9] | 27.5 (4.4),<br>27.2 [24.3 - 29.9] | 52,144 |
| **Environmental and lifestyle factors** |  |  |  |
| Urban environment | 44 401 (85.4%) | 301 (86.0%) | 52,331 |
| Smoking history |  |  | 52,078 |
| Never smoked | 28 213 (54.3%) | 142 (40.6%) | 28,355 |
| Current or ex-smoker | 23 516 (45.7%) | 207 (59.4%) | 23,723 |
| NSAID exposure | 14 877 (28.6%) | 97 (27.7%) | 52,331 |
| **Laboratory tests** |  |  |  |
| C-Reactive Protein (mg/L) | 2.64 (4.45),<br>1.34 [0.66 - 2.80] | 3.67 (5.09),<br>1.95 [0.87 - 4.37] | 50,200 |
| Albumin (g/L) | 45.2 (2.7),<br>45.1 [43.4 - 46.9] | 44.6 (2.8),<br>44.8 [43.1 - 46.2] | 46,952 |
| Leukocyte count (10<sup>9</sup> cells/L) | 6.88 (2.13),<br>6.65 [5.60 - 7.87] | 7.18 (1.92),<br>6.90 [6.00 - 8.11] | 50,938 |
| Haemoglobin (g/dL) | 14.1 (1.3),<br>14.1 [13.3 - 15.0] | 14.1 (1.4),<br>14.1 [13.2 - 15.1] | 50,939 |
| Platelet count (10<sup>9</sup> cells/L) | 252.5 (59.8),<br>248.0 [213.0 - 287.0] | 264.4 (75.7),<br>256.0 [217.9 - 304.7] | 50,939 |
| Triglycerides (mmol/L) | 1.74 (1.02),<br>1.48 [1.05 - 2.14] | 1.93 (1.11),<br>1.65 [1.15 - 2.30] | 50,284 |
| Cholesterol (mmol/L) | 5.66 (1.16),<br>5.62 [4.86 - 6.40] | 5.57 (1.15),<br>5.56 [4.72 - 6.38] | 50,319 |
| Creatinine (μmol/L) | 73.0 (20.2),<br>70.7 [61.5 - 81.4] | 74.2 (20.5),<br>71.0 [60.6 - 82.3] | 50,303 |
| Vitamin D (nmol/L) | 48.5 (21.1),<br>46.7 [32.3 - 62.3] | 48.7 (21.7),<br>46.5 [31.7 - 63.9] | 48,520 |

### Results from feature selection

The top 20 proteins from each model, ranked by mean absolute SHAP values, are summarised in Figure 3. Feature importance plots inform overall importance (Figure 3a), where the most important proteins have the highest absolute SHAP value, while SHAP beeswarm plots inform directionality of effect (Figure 3b): one point represents one participant in the training data, the colour gradient indicates the feature value (i.e. the level of expression of the protein), and the X-axis represents the raw SHAP value of that protein for that participant.

**Figure 3.**
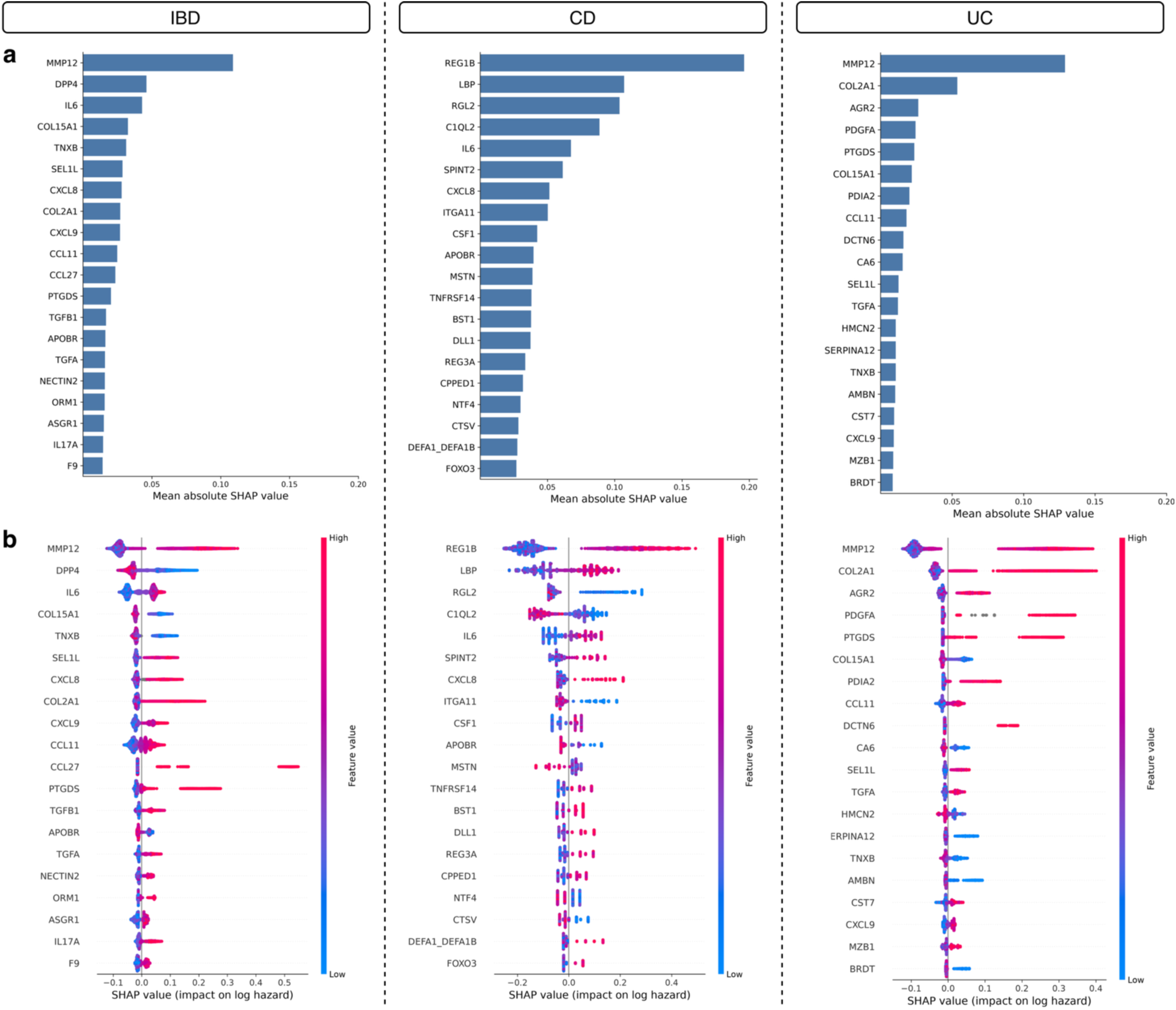
Feature importance and beeswarm plots showing top proteomic machine learning features. **a)** The feature importance plots rank proteins by the mean absolute SHAP value, where the SHAP value represents the marginal contribution of that protein to the overall prediction and a higher value indicates greater importance. **b)** The beeswarm plots represent individual predictions as a scatterplot, where each dot represents one participant, the distribution along the x-axis represents the SHAP values, the colour of the dots represent the feature value where red is a higher value and blue a lower value, and distribution along the y-axis represents clustering around that area.

#### IBD

In the IBD model, MMP12, DPP4 and IL6 had a mean absolute SHAP value of more than 0.05, after which there was a slow taper in the SHAP values up to the twentieth protein, indicating that the lower ranked proteins contribute only marginally to the overall prediction in isolation of the other proteins. At a higher level of protein expression, noted by red points, four proteins (DPP4, COL15A1, TNXB and APOBR) had a negative SHAP value, showing a negative association (i.e. are protective) with developing IBD. The other 16 proteins showed a positive association (i.e. are hazardous), with an increased risk at higher levels of expression.

#### UC

Similar to the IBD model, MMP12 and COL2A1 had a mean absolute SHAP value of 0.05 or more in the UC model, followed by a similar slow drop. In the beeswarm plot, 7 proteins (COL15A1, CA6, HMCN2, SERPINA12, TNXB, AMBN and BRDT) showed a potentially protective effect on developing UC. 9 proteins are shared by the UC and IBD models (MMP12, COL2A1, AGR2, SEL1L, CCL11, PTGDS, TGFA, COL15A1 and TNXB), and show the same association as in IBD.

#### CD

8 proteins (REG1B, LBP, RGL2, C1QL2, IL6, SPINT2, CXCL8, ITGA11) in the CD model had a mean absolute SHAP value of 0.05 or more, compared to 2 proteins in the UC and IBD models, again followed by a slow taper.

In the SHAP beeswarm plot, there were 7 proteins (RGL2, C1QL2, ITGA11, APOBR, MSTN, NTF4 and CTSV) showing a potentially protective association, while the other proteins showed a positive association with developing CD. Three proteins (APOBR, IL6, CXCL8) are shared by the IBD model, and no proteins are shared by the UC and CD models. The most likely explanation is that the IBD model chooses proteins that are predictive of both UC and CD, whereas in their separate models the objective is to find features that are predictive of one subtype but not of the other.

### Statistical analysis using Cox Models

All clinical models except for CD had poor ability to predict diagnosis at 14 years, with a 14-year AUROC of 0.59 for IBD, 0.58 for UC and 0.63 for CD, and a Harrell’s C-index of 0.596 for both the IBD and UC models and 0.62 for the CD model.

In comparison, the proteomics models were better at predicting diagnosis at 7 years and retained this performance at 14 years before diagnosis. The UC proteomics model had the best discrimination when comparing C-indexes (UC: 0.680, IBD: 0.648, CD: 0.606). This trend is further supported by the time-dependent ROC curves, where the UC model performed the best at 14 years with an AUROC of 0.70, compared to an AUROC of 0.66 for IBD and 0.61 for CD. All three models performed similarly at 7 years (AUROC = 0.69 for IBD and UC and 0.70 for CD), better than their respective AUROCs at 14 years.

The combined Cox model assessed whether the clinical features and proteomic features offer similar or complementary predictive information. Performance did not improve across all three combined Cox models compared to the proteomic Cox models, indicating that the full predictive information is likely captured in the proteomic features.

Model performance for the clinical, proteomic and combined multivariate Cox models is summarised in Table 2, showing the C-index and 7-and 14-year time-dependent AUROC across the three disease groups.

**Table 2.** Summary of model performance of the three Cox models (clinical, proteomics and combined) in predicting incident IBD, CD and UC. Proteomics models outperform clinical models in IBD and UC, and the combined models show only a marginal improvement over proteomics models.

| Prediction model | Training dataset (80%) |  |  | Test dataset (20%) |  |  |
| --- | --- | --- | --- | --- | --- | --- |
|  | AUROC at<br>7 years (95% CI) | AUROC at<br>14 years (95% CI) | C-index | AUROC at<br>7 years (95% CI) | AUROC at<br>14 years (95% CI) | C-index |
| **IBD** (cases in training dataset: n=280, cases in test dataset: n=70) |  |  |  |  |  |  |
| Clinical model | 0.67 (0.62 - 0.71) | 0.65 (0.62 - 0.68) | 0.645 | 0.64 (0.55 - 0.74) | 0.59 (0.52 - 0.66) | 0.596 |
| Proteomics model (Top 20 proteins) | 0.76 (0.75 - 0.76) | 0.73 (0.72 - 0.73) | 0.720 | 0.69 (0.59 - 0.79) | 0.66 (0.59 - 0.72) | 0.648 |
| Combined model (clinical + proteomics) | 0.76 (0.75 - 0.77) | 0.73 (0.72 - 0.74) | 0.728 | 0.69 (0.59 - 0.79) | 0.65 (0.59 - 0.72) | 0.651 |
| **CD** (cases in training dataset: n=93, cases in test dataset: n=23) |  |  |  |  |  |  |
| Clinical model | 0.69 (0.61 - 0.77) | 0.69 (0.64 - 0.74) | 0.686 | 0.63 (0.45 - 0.81) | 0.63 (0.52 - 0.74) | 0.620 |
| Proteomics model (Top 20 proteins) | 0.79 (0.77 - 0.80) | 0.81 (0.80 - 0.82) | 0.799 | 0.70 (0.50 - 0.90) | 0.61 (0.48 - 0.75) | 0.606 |
| Combined model (clinical + proteomics) | 0.81 (0.79 - 0.83) | 0.82 (0.81 - 0.82) | 0.807 | 0.66 (0.42 - 0.91) | 0.61 (0.47 - 0.75) | 0.609 |
| **UC** (cases in training dataset: n=184, cases in test dataset: n=47) |  |  |  |  |  |  |
| Clinical model | 0.68 (0.63 - 0.73) | 0.66 (0.62 - 0.70) | 0.651 | 0.63 (0.52 - 0.74) | 0.58 (0.49 - 0.67) | 0.596 |
| Proteomics model (Top 20 proteins) | 0.75 (0.75 - 0.76) | 0.74 (0.74 - 0.75) | 0.730 | 0.69 (0.59 - 0.80) | 0.70 (0.62 - 0.78) | 0.680 |
| Combined model (clinical + proteomics) | 0.76 (0.76 - 0.77) | 0.76 (0.75 - 0.76) | 0.747 | 0.68 (0.58 - 0.79) | 0.70 (0.62 - 0.78) | 0.683 |

On the time-dependent AUROC plots (Figure 4), performance in all three models is best in the shorter term, after which it stabilises. The performance spike observed at 16 years across all three models is likely an artefact of heavy censoring between 14 and 16 years (>90% of the cohort censored), which can artificially inflate performance, and we therefore excluded the 16-year timepoint from the discussion.

**Figure 4.**
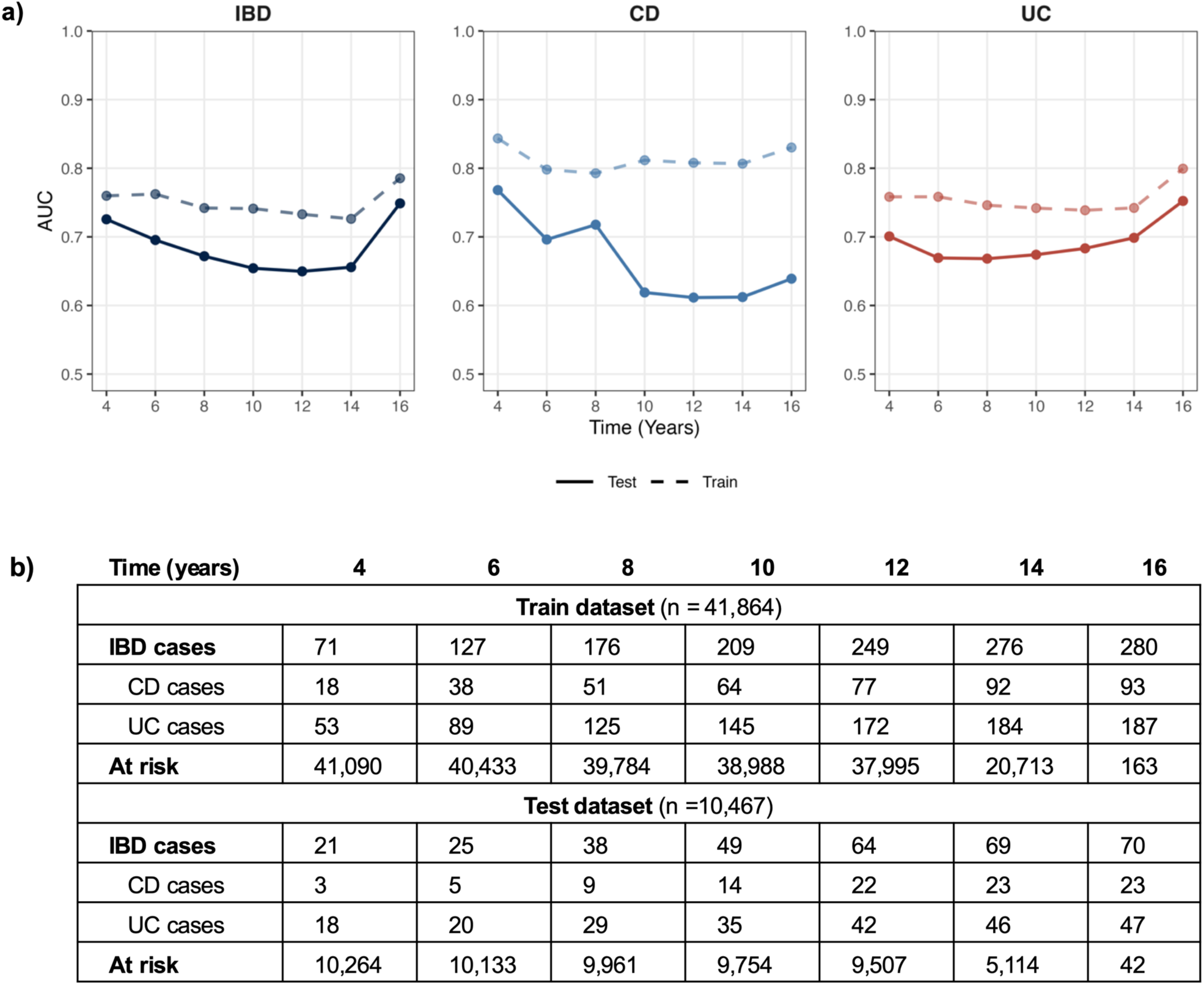
Time-dependent area under receiver operator curve (AUROC) plot for the proteomics Cox models. **a)** At each timepoint, the AUROC reflects the model’s ability to distinguish cases from those still at risk. To account for censoring, participants were weighted by their probability of developing IBD, estimated using inverse-probability-of-censoring weights derived from a Kaplan–Meier survival curve. **b)** The table shows the total number of cases for each disease group at each timepoint, with the number at risk those that have not experienced an event of interest at that timepoint.

#### Hazard ratios and model outputs

Figure 5 visualises the hazard ratios (HR) of the predictors across the three models for each disease group. Predictors where the 95% confidence interval (CI) crosses the 1-line were non-significant. The values of the hazard ratios and significance levels are available in the Supplementary Table S2.

**Figure 5.**
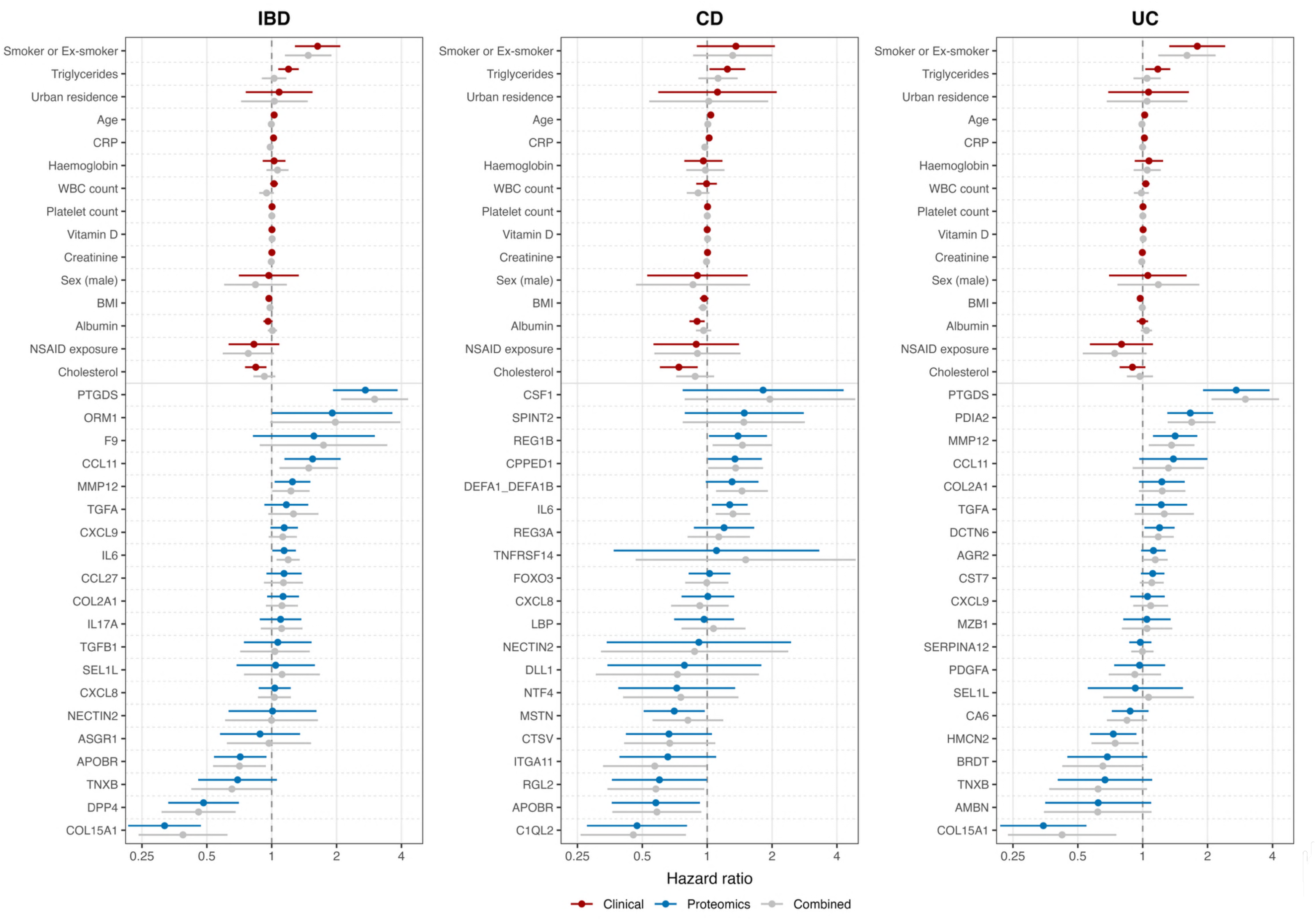
Forest plot showing the hazard ratios (dots) and 95% confidence intervals (horizontal line) for the clinical, proteomics and combined Cox models.

### Clinical models

The clinical Cox models for IBD, UC and CD included 15 predictors identified in existing literature. In all three models, smoking had the highest hazard ratio and reached significance in IBD and UC (IBD: HR = 1.63, p < 0.001, UC: HR = 1.79, p < 0.001, CD: HR = 1.36, p = 0.15). This is unexpected, as smoking is considered a protective factor in UC, but it has been documented that risk can increase paradoxically after smoking cessation^21^. To assess whether smoking cessation contributed to this, smokers and ex-smokers were split into two separate predictors, and in all three models both still had hazard ratios of more than 1.

Triglyceride levels were hazardous in all three models with statistical significance (IBD: HR = 1.20, p = 0.0013; UC: HR = 1.18, p = 0.018; CD: HR = 1.24, p = 0.027), and cholesterol levels were protective in all three models, reaching statistical significance in IBD and CD (IBD: HR = 0.84, p = 0.0035, UC: HR = 0.90, p = 0.12, CD: HR = 0.74, p = 0.0033). Albumin levels were protective only in CD (HR = 0.90, p = 0.091) and had no effect in IBD and UC.

Age and platelet count were significant predictors across all three models, and CRP and body mass index (BMI) in the IBD model only. Their hazard ratios were close to 1, however, indicating weak associations and limited contribution to the overall risk prediction.

### Proteomics Cox models

In the IBD model, 5 proteins were statistically significant risk factors (PTGDS, ORM1, CCL11, MMP12 and IL6), and 10 proteins had associated risk but did not reach statistical significance (F9, TGFA, CXCL9, CCL27, COL2A1, IL17A, TGFB1, SEL1L and CXCL8). 3 proteins were associated with a statistically significant protective effect (APOBR, DPP4 and COL15A1), with TNXB having a protective effect but not achieving significance. These were all in accordance with the associations that the SHAP beeswarm plot identified.

Similarly in CD, all significant proteins exhibited the same directional association as their SHAP values. 3 proteins were statistically significant risk factors (REG1B, CPPED1 and IL6), and 4 proteins (MSTN, RGL2, APOBR and C1QL2) were associated with a significant protective effect of developing CD. The UC model had the fewest statistically significant proteins: PTGDS, PDIA2 and MMP12 were all associated with a significant risk, and HMCN2 and COL15A1 were both protectively associated with developing UC.

### Combined Cox Model

Smoking remained a statistically significant risk factor in both IBD and UC, while all other clinical features lost significance. Therefore, with the exception of smoking, the clinically attributable variance from the demographic and biochemical features appears to be fully captured by the proteomics features.

Most proteins retained their significance in all the combined Cox models. The exceptions were ORM1, which lost significance in the combined IBD model, DEFA1 and ITGA11, which became significant in the combined CD model, and AGR2, which became significant in the combined UC model.

## Discussion

This study of 52,331 UKB participants shows that combining machine learning with statistical modelling identifies proteins associated with incident IBD and predicts it up to 14 years before diagnosis, while outperforming known clinical risk factors.

Furthermore, our results show that machine-learning feature selection followed by multivariate Cox prediction compares to the reverse approach of Feng et al.^18^ and Zhang et al.^17^, who used univariate Cox models for feature selection and machine learning for prediction. At the median time to diagnosis (7 years), our IBD proteomics model showed similar performance to Zhang et al. (AUROC 0.69 vs 0.68), and retained its predictive ability up to 14 years before diagnosis with an AUROC of 0.64. Notably, our UC model performed better than that of Zhang et al. at 14 years before diagnosis (AUROC 0.70 vs 0.61), while our CD model performed marginally worse than both Zhang et al. and Feng et al. (AUROC 0.61 vs 0.62 and 0.76 respectively)^17,18^. However, the results of Feng et al. may be over-optimistic, as their use of class balancing likely inflated performance relative to the true event distribution encountered in the real world.

In all three disease groups, our clinical models performed better than theirs, a reflection of the careful selection of relevant clinical predictors from literature. Interestingly, smoking was hazardous for developing IBD and UC, in contrast to existing literature, and was the only clinical predictor to retain its significance in the combined Cox model. This could be an effect seen only in this dataset, but it does suggest that smoking is an important modifiable risk factor that should receive attention in further research.

Our IBD model shared 7 of the 10 proteins identified by Zhang et al., consistent with the two IBD models performing similarly. Notably, our CD model selected a largely different set of proteins yet achieved comparable performance, sharing 2 proteins with Zhang et al.’s model and 3 with Feng et al.’s model, which suggests that no single protein signature is uniquely predictive for CD. By contrast, our UC model shared only two proteins with Zhang et al.’s; here the smaller overlap coincided with superior performance, which could mean that our model selected more predictive proteins for UC^17^. All shared proteins can be found in Supplementary Table S3 online.

Taken together, the variability in the proteins selected across IBD, CD and UC, despite similar predictive performance within the same population, suggests that many proteins are associated with these conditions and that those which prove most predictive may depend heavily on the specific dataset distribution, such as a specific train/test split. Further research could test this pipeline across multiple train/test splits to identify proteins that are consistently predictive.

### Protein Analysis

We used the online databases UniProt^22^, ProteinAtlas^23^, String-DB^24^ and existing literature to understand the function of the proteins selected by the models. Most of the proteins are already associated with IBD, and serve roles in known inflammatory pathways, extracellular matrix (ECM) reorganisation, and lipid metabolism, with 14 proteins across the 3 models not being directly linkable to one of these functions. This provides reassurance that the models select proteins that are meaningfully associated with disease. Proteins that are not previously associated with IBD are discussed in detail in the Supplementary Material.

#### Inflammatory proteins

Three inflammatory pathways are enriched in the identified proteins, the Interleukin-4/Interleukin-13 (IL-4/IL-13) pathway, the IL-23/IL-17 pathway, and the IL-10 pathway. The complex signalling between the identified proteins and other inflammatory proteins are shown in Figure 6 and explained below.

**Figure 6.**
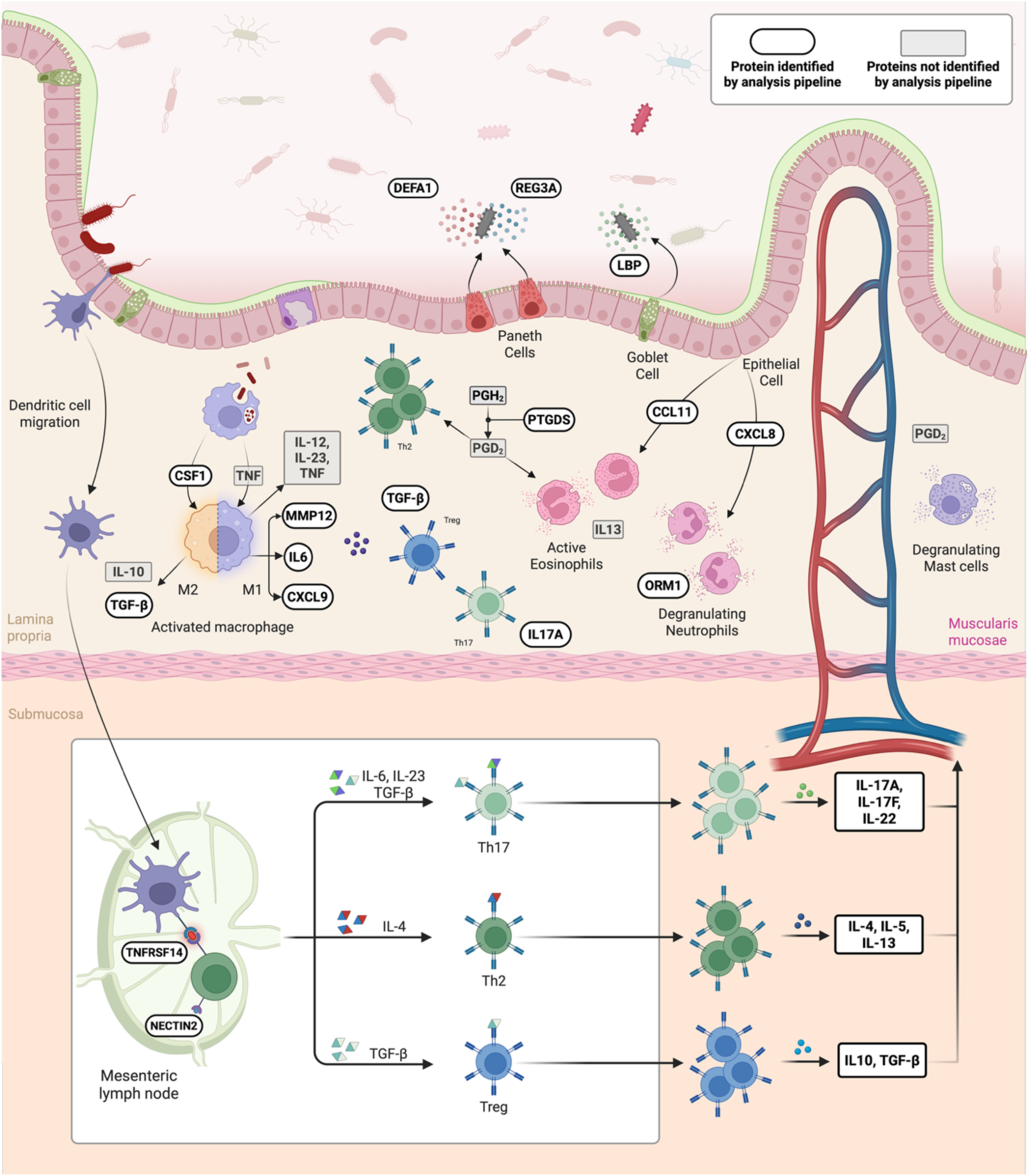
Proteins identified by the pipeline mapped to their known biological functions in the intestinal mucosa. It is proposed that through a complex interaction between various proteins in the identified inflammation pathways, a deranged immune response is maintained in response to pathological bacteria. The inflammatory pathways identified by my models were the IL-17/IL-23 pathway with a predominant Th17 response through IL-6 and TGF-B, the IL-4/IL-13 pathway with a predominant Th2 response, and IL-10 pathway with a predominant Treg response through TGF-B. CSF1 promotes the maturation of macrophages into the immunosuppressive M2 subtype. Other proteins play various role in chemoattraction (CCL11, CXCL8, PTGDS), cell-to cell signalling (TNFRSF14, NECTIN2), leukocyte degranulation (ORM1, CCL11, PTGDS), host protection against bacteria (REG3A, DEFA1, LBP) and induction and maintenance of T-cell response (TGF-B, IL17A, IL6, CXCL9, TGF-β, PTGDS). **CCL11**, C-C motif chemokine ligand 11; **CSF1**, colony-stimulating factor 1; **CXCL8/9**, C-X-C motif chemokine ligand 8/9; **DEFA1**, defensin alpha 1; **IL**, interleukin; **LBP**, lipopolysaccharide-binding protein; M1/M2, classically/alternatively activated macrophages; **MMP12**, matrix metalloproteinase 12; **NECTIN2**, nectin cell adhesion molecule 2; **ORM1**, orosomucoid 1; **PGD2**, prostaglandin D2; **PGH2**, prostaglandin H2; **PTGDS**, prostaglandin D2 synthase; **REG3A**, regenerating family member 3 alpha; **TGF-β**, transforming growth factor beta; **Th**, T helper cells; **TNF**, tumour necrosis factor; **TNFRSF14**, tumour necrosis factor receptor superfamily member 14; **Treg**, regulatory T cell.

The IL-4/IL-13 pathway, which drives a Th2-type response, is represented by IL-6, TGFB1, CCL11, IL17A and CXCL8. The proinflammatory IL-23/IL-17 pathway, which promotes a Th17-driven neutrophilic response, is represented by IL6, IL17A and TGFB1. The anti-inflammatory IL-10 pathway, which regulates the other two through regulatory T-cells, involves TGFB1 (TGF-β), FOXO3 and CSF-1. A fuller description of each pathway and its associated proteins is given in the Supplementary Material.

PTGDS, which had the highest hazard ratio in the IBD model, synthesises prostaglandin-D2, which plays an active role in all of the above pathways by promoting both Th2 and M2 differentiation^22,25^. Other proteins that are involved in various functions in the immune system are ORM1^26^, NECTIN2^27^, LBP^28^, DLL1^29^, BST^22^, CCL27^30^, DEFA1^31^, TNFRSF14^22^, CST7^32^ and REG3A^33^.

#### Extracellular matrix proteins

TGFA, COL15A1, TNXB, COL2A1, ITGA11, REG1B and MMP12 are associated with the ECM or tissue repair^22,23,34^, helping to remodel the ECM in response to injury.

MMP12 is released by inflammatory macrophages and could contribute to increased permeability that allows bacteria and inflammatory cells to enter the intestine^14,35,36^. MMP12 is markedly higher in UC compared to CD and has been associated with severe disease^36^, explaining why it was significant in the UC and not in the CD model. COL15A1 is necessary to maintain the normal ECM, and in murine studies of UC its levels decrease in response to inflammation^37^.

#### Lipid metabolism

Five identified proteins are involved in lipid metabolism (APOBR, SERPINA12, CPPED1, MZB1 and DPP4)^22^. Of these, APOBR is the only one with a protective association. APOBR is a receptor for apolipoprotein B-containing lipoproteins, which transport lipids between the gut, liver and peripheral tissues, and reduced APOBR would impair clearance of triglyceride-rich lipoproteins, allowing triglycerides to accumulate and impairing cholesterol synthesis. This is consistent with previous findings^38,39^ and our clinical model, where elevated triglycerides and reduced cholesterol are associated with developing Crohn’s disease.

DPP4 is an enzyme that breaks down peptides such as GLP-1s, inflammatory proteins and cytokines in the bloodstream, and also acts as an important co-signalling protein in T-cell signalling^40^. A recent systematic review^41^ has found that DPP4 levels tend to decrease in active CD, explaining the protective association.

#### Other proteins

14 identified proteins have functions which do not clearly fit into the current understanding of the pathogenesis of IBD, for example MSTN inhibits muscle growth^22^, C1QL2 supports synapse formation in the brain^22^, and DCTN6 is an accessory protein in the dynactin complex^22^. Other proteins include F9, AGR2, BRDT, AMBN, NTF4, CTSV, RGL2, PDIA2, DCTN6, HMCN2 and SPINT2. Their predictive value is unexplained by their functions, and more research is warranted to investigate the link between these proteins and IBD. These are discussed further in the Supplementary Material.

### Strengths and limitations

Through outcome-agnostic methods, we have demonstrated the use of machine learning and statistical models to identify potential protein predictors for IBD and its subtypes from 2,912 proteins. The long follow-up time in the UKB allowed us to demonstrate model performance over a 14-year time horizon, indicating a strength in detecting proteins associated with preclinical rather than purely undiagnosed disease. The IBD models selected mostly proteins that are already associated with IBD, confirming previous findings in literature through different methods and demonstrating that this pipeline identified true biological signal, while the UC and CD models also selected some proteins that are not directly associated with disease and could be explored in future research.

However, our approach does have some limitations. The UKB has a predominantly white ethnicity and only includes residents in the United Kingdom, limiting its generalisability to other regions, as genetic and environmental factors play an important role in disease pathogenesis. The UKB also only enrolled participants between the ages of 40-69 years, whereas the peak age of diagnosis is usually between 20-30 for CD and 30-40 for UC^2^, so cases of IBD in this study may not be representative of the full spectrum of disease. Future studies should seek to validate these findings in external and geographically distinct datasets across the full disease spectrum.

A further limitation is the potential risk of overfitting to noise during feature selection, attributed to a very low event rate and a high number of proteomic features. This is mitigated through XGBoost’s regularisation and subsampling hyperparameters, and the fact that most proteins were previously associated with IBD provides reassurance that these findings could reflect true biological associations. Despite this, the models’ performance is still suboptimal, limiting their use in clinical practice. Some proteins could have a non-linear effect, and to keep our Cox models parsimonious, we refrained from assessing non-linearity and adding spline terms, which could have contributed to the suboptimal performance.

Finally, our models were developed from a single cross-sectional proteomic dataset, as the UK Biobank provides only one measurement per participant. A single time point cannot capture the dynamic, evolving nature of inflammation in UC and CD, and future research applying proteomics at multiple time points before clinical diagnosis could characterise this dynamic behaviour and reveal how these proteins change over time.

## Conclusion

As the global burden of IBD grows, the focus is shifting from diagnosing and treating the disease towards predicting and preventing it. Proteomics offers insight into disease pathogenesis that could support prediction, risk stratification and, ultimately, prevention. In this study, we demonstrated an analysis pipeline that combines machine learning for feature selection with Cox modelling to identify proteins associated with incident IBD, CD and UC. Across all three outcomes, our proteomic models predicted disease up to 14 years before diagnosis, performed better than clinical risk factors, and outperformed models reported in the existing literature. Most of the selected proteins had previously been linked to IBD, indicating that the pipeline detects biologically meaningful signal.

These findings have several implications for clinical practice and research. With further development and external validation, the identified proteins could be incorporated into risk prediction tools to detect preclinical IBD and shorten the diagnostic pathway, for example by applying a targeted protein panel to individuals at high genetic risk. The proteins could also guide targeted research into IBD pathophysiology and be investigated as potential drug targets. Finally, the pipeline could be applied to other chronic conditions, providing a framework for identifying candidate proteins in disease areas that remain poorly understood. However, external validation in independent cohorts is now needed before these findings can be translated into practice.

## Methods

### Study design and participants

This observational cohort study was performed on a subset of participants in the UKB who had baseline proteomics data available, without prevalent IBD.

The UKB is a population-based prospective cohort study, which enrolled nearly half a million participants aged 40-69 years between 2006 – 2010 from 22 assessment centres across England, Wales and Scotland^42^. Baseline information spanning multiple domains was collected during the initial visit through questionnaires, verbal interviews, physical examinations, and collection of biological samples. The study has ethical approval from the National Health Service Northwest Multicentre Research Ethics Committee (06/MRE08/65) and was conducted under UKB protocol number 83801. All participants provided written consent prior to enrolment, and all methods were performed in accordance with the relevant guidelines and regulations.

The UKB contains demographic, lifestyle and self-reported health data, genomic data, proteomic data, primary care and hospital inpatient data through linkage with healthcare records, and cancer and death data through registry linkage, providing a rich measurement of participants’ overall health trajectory.

### The UKB Pharma Proteomics Project

The UKB Pharma Proteomics Project (UKB-PPP) ran between 2021 and 2022. Using blood specimens collected at the baseline visit for proteomic profiling, up to 2,923 plasma proteins of 54,219 participants in the UKB were analysed using the antibody-based Olink Explore 3072 proximity extension assay^43^.

The UKB-PPP cohort was derived through random selection of 46,595 UKB participants at the baseline visit, 6,376 participants selected by the UKB-PPP consortium members, and 1,268 participants attending the COVID-19 repeat-imaging study. The prevalence of UC and CD was not significantly enriched in the UKB-PPP cohort compared to the UKB cohort (UC: 1.32% vs 1.25%, CD: 0.72% vs 0.65%)^43^.

The UKB-PPP consortium ensured high data quality by implementing comprehensive internal quality control measures described in detail elsewhere^43^. Protein levels were expressed as normalised protein expression (NPX) values representing the relative abundance of a protein on a logarithmic scale around a median of zero, rather than its absolute concentration.

### Definition of prevalent and incident cases

Cases were identified using the International Classification of Diseases (ICD) codes for CD (ICD-9: 555.X; ICD-10: K50.X), and UC (ICD-9: 556.X; ICD-10: K51.X). ICD-9 codes became obsolete in 1996, thus all occurrences were considered to be prevalent cases. UKB data fields 131626 and 131628 were used to determine the date of first occurrence for ICD-10 codes K50.X and K51.X respectively, representing their date of first occurrence in primary care records, inpatient hospital records, death registry data or self-reported diagnoses.

Prevalent cases of IBD were defined as having a diagnosis of either CD or UC before the date of the baseline visit to the UKB assessment centre, and incident cases after this date. The time to diagnosis was calculated as the number of years from the baseline visit until the first occurrence of UC or CD. In non-cases, the follow-up time for censoring was calculated as the number of years until the earliest of the following: death from any cause, loss of follow-up, or the end of linkage with hospital inpatient records in the UKB (31 March 2023 in England, 31 August 2022 in Scotland and 31 May 2022 in Wales).

The final study population was split into a training (80%) and a test set (20%). To ensure equal distribution of the outcomes, the training and test sets were balanced for both CD and UC cases, resulting in a balanced split for total IBD cases which remained identical to their incidence rates in the full dataset.

### Competing risks

In subgroup analyses for UC and CD, the two subtypes were handled as competing risks, as a diagnosis of one IBD subtype changes the subsequent risk of developing the other, and would violate the assumption of non-informative censoring.

We elected to use a cause-specific proportional hazard model, which is most commonly used in etiological studies to describe the underlying causes of disease^44,45^, rather than a sub-distribution hazard model (known as a Fine-Gray model), which is used in epidemiology to model the incidence of disease in a given population^44,45^. We censored the competing IBD subtype at the time of diagnosis, and those participants no longer form part of the population at risk. In cases where both diagnoses were present, the first diagnosis was taken as the outcome.

### The analysis pipeline

#### Overview

Liu et al. described a pipeline that combines both ML and conventional statistical methods for survival analyses^46^. In their approach, feature selection is performed by using XGBoost, after which the most predictive features are used as covariates in a conventional Cox model. Fieggen et al. adapted this pipeline to predict the incidence of multiple myeloma in the UKB using proteomics^47^, forming the basis of this analysis.

Firstly, traditional risk factors were identified from existing literature. Secondly, an XGBoost model was developed using 2,912 protein predictors in the UKB-PPP, and SHAP values were used to select the top 20 most important proteins from the XGBoost model. Finally, three different Cox models were developed: a clinical Cox model with only traditional risk factors as covariates, a proteomics Cox model using only the identified top 20 proteins, and a combined Cox model that included both the clinical and protein predictors. This pipeline was applied to IBD as well as the two subtypes CD and UC, resulting in three different disease group analyses through the same pipeline (see Figure 1 on pg. 4).

#### Identifying traditional clinical risk factors

In the absence of validated clinical risk prediction models, we identified 15 risk factors across demographic features, environmental exposures, and laboratory tests that have been associated with developing UC and CD and are available in the UKB. These were chosen if they were a known risk factor of developing IBD, a biochemical test used routinely to investigate suspected IBD, or were found to be associated with IBD in multiple studies.

Among demographic features, age, sex and BMI were chosen as predictors^2,48^. Ethnicity was not used as a predictor as IBD occurs equally among different populations^2^. The chosen environmental exposures were urban living, smoking, and exposure to non-steroidal anti-inflammatory drugs (NSAIDs), which are all associated with an increased risk of developing IBD^21,49,50^.

The laboratory tests included haemoglobin, white cell count, platelet count, creatinine and CRP^8,9^. Notably, faecal calprotectin, the most sensitive and specific test for IBD^8^, is unavailable in the UKB and therefore could not be included. Vitamin D^21^ and triglyceride levels^38,39^ are associated with an increased risk of developing IBD, and albumin^51^ and cholesterol levels^38,39^ decrease with severe CD, therefore these were included in our analysis.

#### Data preprocessing

Minimal data pre-processing was required as the UKB already has rigorous data processing standards. Smokers and ex-smokers were combined into one smoking category. NSAID exposure was derived from the self-reported medication field and any reported NSAID use was taken as positive (medication codes are available in Supplementary Table S1 online). Urban living was identified from the home population density field, using the classification already provided in the UKB.

Within this study, we excluded 14 proteins that were missing more than 20% of their values, leaving 2,912 proteins for analysis. Proteomic values were not pre-processed as they already underwent rigorous quality checks and standardisation^43^.

#### Proteomic feature selection using XGBoost

XGBoost, an ensemble tree boosting algorithm, was selected for feature selection because it can handle non-linear relationships between correlated predictors as well as between predictors and the outcome^52^, natively handles missingness, and can rank numerous features by importance using explainability techniques such as SHAP values^53^. An added benefit is its superior performance with minimal finetuning when compared to other conventional and deep-learning ML algorithms, especially when applied to large tabular datasets with a big feature space^54,55^.

### XGBoost Model training

The training dataset was used to train three separate XGBoost models for IBD, CD and UC, with “cox-loss” as the loss function, which aims to reduce the negative partial log-likelihood of the Cox proportional hazard model. Missingness was regarded as informative and assigned a default direction in each split based on which optimised the loss function the most. Overfitting was assessed by calculating Harrell’s C-index on both the training and the held-out test datasets.

The hyperparameters were tuned on the three models separately using grid search with five-fold cross-validation, selecting the hyperparameters that reduced the cox-loss function the most in each disease group. The hyperparameters and full tuning strategy are available in the Supplementary Material.

The IBD model and the UC model showed moderate overfitting with a training and test C-index of 0.882/0.644 and 0.906/0.661 respectively. The CD model had the worst overfitting with a training C-index of 0.973 and test C-index of 0.608. This is likely due to severe class imbalance in the dataset due to the low event rate (IBD: 0.6%, UC: 0.4%, CD: 0.2%). No weighting was applied to cases, and the ratio between cases and healthy controls was kept unbalanced to reflect model performance in real-world settings.

SHAP values were used for feature importance since they offer the most stability when compared to other methods^56^, which is especially important in datasets where event rates are rare and minor changes in the dataset can occur easily (e.g. participants dropping out)^53^. A feature’s SHAP value is calculated from principles of cooperative game theory as the average of its individual contribution to the prediction in different feature combinations, weighted by the probability that that combination will occur^53^. Feature importance was calculated as the mean of the absolute values of each feature’s SHAP value per participant, proteins were ranked accordingly, and the top 20 proteins were selected for further evaluation in the statistical analyses.

#### Statistical analyses

Conventional statistical methods such as the Cox proportional hazards model require care both in selecting non-correlated features and in handling missingness. Correlation was assessed using the pairwise correlations (*r*) between the different types of features in the training dataset: Spearman’s rank coefficients for pairs of numeric features, point-biserial correlations between numerical and categorical features, and Cramer’s V correlation heatmap for pairs of categorical features. No features were found to be highly correlated (*r* > 0.9).

Missingness was assumed to be at random, and was handled by using mode imputation for categorical variables, and multiple imputation for continuous variables through 5 iterations per round over ten imputation rounds using the *mice* package in R^57^.

Thereafter, three Cox proportional hazards models were constructed per disease group: a clinical Cox model, comprising the identified clinical predictors, a proteomic Cox model with the top 20 proteins as predictors, and a combined Cox model using both the clinical and proteomic predictors.

The proportional hazards assumption of the Cox models was assessed visually using scaled Schoenfeld residual plots, and multi-collinearity was assessed using variance inflation factors (VIF), where any VIF < 10 was deemed acceptable. The statistical tests were two-tailed using a significance level of 5%.

### Evaluating model performance

Model performance was assessed using Harrell’s C-index, a time-independent measure of the model’s ability to rank participants in order of event time based on their risk score, and the time-dependent AUROC^58^.

The time-dependent AUROC was used to compare model performance at different follow-up times, while incorporating appropriate handling of censoring *through inverse probability of censoring weighting. Using the timeROC package* in R, the AUROC was calculated at 4, 6, 8, 10, 12, 14 and 16 years, discriminating IBD events at each time point from the population still at risk (uncensored).

The Brier score is often used as a measure of calibration in survival analyses, but is highly sensitive to class imbalance, and would be overly optimistic if used in this analysis^59^.

## Software used

Proteomic feature selection was performed using XGBoost (2.1.4) and SHAP (0.51.0) run on Python (3.11), and statistical analysis was done in R (4.3.2) using the *mice* package (3.19.0) for multiple imputation, and the *survival* package (3.8.6) to construct Cox models. Other packages included numpy (2.4.6), pandas (3.0.3), matplotlib (3.10.9), scikit-learn (1.8.0) and scikit-survival (0.27.0) in Python and ggplot2 (4.0.3), timeROC (0.4.1) and tidyverse (2.0.0) in R. The code was compiled using Claude Code by Anthropic. The TRIPOD checklist was used as the reporting guideline for predictive models (see Supplementary Material for full checklist)^60^.

## Use of generative AI

Generative AI was used for coding through Anthropic’s Claude Code interface using a combination of Opus 4.8 and Sonnet 4.6, depending on the complexity of the task. Prompts were written with explicit instructions of what the objective of the code was, including where possible which packages to use. The code was reviewed manually before running, and the outputs reviewed carefully.

## Acknowledgements

This research was conducted using the UK Biobank Resource under application number 83801. This work used data that were provided by patients and collected by the National Health Service (NHS) as part of their care and support. The authors thank the participants of the UK Biobank study without whom this research would not have been possible. For computation, this study used the Oxford Biomedical Research Computing facility, a joint development between the Wellcome Centre for Human Genetics and the Big Data Institute supported by Health Data Research UK and the National Institute of Health Research (NIHR) Oxford Biomedical Research Centre (BRC).

DAC was funded by an NIHR Research Professorship (NIHR302440); a Royal Academy of Engineering Research Chair; and the InnoHK Hong Kong Centre for Cerebro-cardiovascular Engineering (COCHE); and was supported by the National Institute for Health Research (NIHR) Oxford Biomedical Research Centre (BRC) and the Pandemic Sciences Institute at the University of Oxford. The views expressed are those of the author(s) and not necessarily those of the NHS, the NIHR or the Department of Health.

We would like to acknowledge the use of generative AI during this study. Generative AI was used for coding through Anthropic’s Claude Code interface using a combination of Opus 4.8 and Sonnet 4.6, depending on the complexity of the task. Prompts were written with explicit instructions of what the objective of the code was, including where possible which packages to use. The code was reviewed manually before running, and the outputs reviewed carefully.

The figures were made in PowerPoint (Figure 1), Draw.io (Figure 2), Python v3.11 (Figure 3) and RStudio v4.3.1 (Figure 4 - 5); for the latter two, the code was written with the help of generative AI as outlined above. Figure 6 was made manually using BioRender.com, using a pre-existing template for inflammatory proteins in IBD. Appropriate licences have been obtained before publication.

## Author contributions

T.E.N., L.C., A.T. and A.P. designed the research; T.E.N., J.F. and B. M. S. analysed the data; T.E.N. drafted the manuscript; and all authors revised and edited the final manuscript.

## Data availability

The data used in this study were obtained from the UK Biobank and are available via application directly to the UK Biobank (https://www.ukbiobank.ac.uk).

