## Supplementary Material for "Serum proteins predict incident inflammatory bowel disease 14 years before diagnosis: a machine-learning and Cox modelling in the UK Biobank"

Tobias Erich Niebuhr[1], Joshua Fieggen[2], Bradley Max Segal[2], Anuj Patel[3], David Clifton[2,3,4], Lei Clifton[1,2]*

[1] Nuffield Department of Primary Care Health Sciences, University of Oxford, Radcliffe Primary Care Building, Radcliffe Observatory Quarter, Woodstock Road, Oxford OX2 6GG, United Kingdom

[2] Computational Health Informatics (CHI) Lab, Institute of Biomedical Engineering, Department of Engineering Science, University of Oxford, Old Road Campus Research Building, Headington, Oxford OX3 7DQ, United Kingdom

[3] Mirae AI, Oxford, United Kingdom (A UK limited company)

[4] OSCAR (Oxford Suzhou Centre for Advanced Research), University of Oxford, Suzhou 215123, Jiangsu, China

*** Corresponding Author:**, Nuffield Department of Primary Care, University of Oxford, Radcliffe Primary Care Building, Radcliffe Observatory Quarter, Woodstock Road, Oxford, OX2 6GG, United Kingdom

### Supplementary Methods – Additional methodological detail

XGBoost is a form of ensemble tree boosting algorithm in which the prediction scores of each decision tree are summed up to get a final prediction score for further downstream tasks such as classification and regression. XGBoost adds an objective during training in the form of a loss function, allowing the model to optimize each decision tree by minimizing the loss function of the preceding decision trees, after which the final prediction score is calculated using the weighted sum of each tree's output.

XGBoost was selected for feature selection compared to other ML algorithms for a few reasons. XGBoost can handle non-linear relationships between correlated predictors as well as between predictors and the outcome.[1] XGBoost is also able to natively handle missingness by assigning a default direction for missing features at each split based on the gain. Lastly, XGBoost has the ability to rank numerous features by importance using explanability techniques such as SHAP values.[2] An added benefit is the superior performance of XGBoost models with minimal finetuning when compared to other conventional and deep-learning ML algorithms, especially when applied to large tabular datasets with a big feature space.[3,4]

Model performance was assessed using Harrell's C-index and the time-dependent Area under the Receiver Operating Characteristic Curve (AUROC). Harrell's C-index is a time-independent measure of model performance, assessing the model's ability to rank participants in order of event time based on their risk score. Events are paired with non-events if the follow-up time of an event is smaller than that of a non-event. If the event has a larger predicted risk than the non-event the pair is concordant, and the C-index is calculated as the ratio of concordant pairs over all pairs.[5]

In contrast, the time-dependent AUROC is used to compare model performance at different follow up times, while incorporating appropriate handling of censoring. Using the package *timeroc* in R, the AUROC was calculated at 4, 6, 8, 10, 12, 14 and 16 years. At each time point, the sensitivity and specificity of the model is calculated at different predicted risk cutoffs, after which the AUROC is calculated. The population still at risk (uncensored) are treated as non-events and discriminated from IBD events at that timepoint. All censored participants are accounted for using inverse probability of censoring weighting, which upweights remaining participants with a similar probability of developing IBD.

### Supplementary Methods – Hyperparameter tuning strategy

The hyperparameters were tuned separately for the IBD, CD and UC XGBoost models through the same process. Grid search was done with a 5-fold cross-validation in the training dataset, the best combination of hyperparameters were chosen that reduced the negative cox-loss function the most in the validation set while having the best C-index. Optimal performance was prioritised above the reduction in the loss function, and in cases where the loss function was equivocal (within 0.05 of each other) the parameter with the highest C-index was chosen.

The grid search was done across 5 stages, and the best hyperparameters were carried forward to the next stage until all hyperparameters were tuned. For stage 1, max depth and minimum child weight were searched, where max depth ∈ [1, 2, 3] and min child weight ∈ [1, 2, 3, 4, 5]. Next, gamma ∈ [0.0, 0.2, 0.4, 0.6, 0.8, 1.0] was searched, followed by subsample and column sample by tree both ∈ [0.5, 0.6, 0.7, 0.8, 0.9]. The regularisation parameters lambda (λ) and alpha (α) were searched in λ ∈ [0.1, 0.5, 1.0, 2.0, 5.0, 10.0, 20.0] and α ∈ [0.0, 0.1, 0.5, 1.0]. Lastly, the learning rate was searched across learning rate ∈ [0.005, 0.01, 0.05, 0.1]. Early stopping was enabled during tuning and the median number of trees stopped at across all 5 folds was used as the maximum number of trees for the final models.

The selected hyperparameters for each model were:

| **Hyperparameter** | **IBD** | **CD** | **UC** |
|---|---|---|---|
| Learning rate | 0.01 | 0.1 | 0.01 |
| Maximum depth | 2 | 2 | 2 |
| Subsample | 0.8 | 0.9 | 0.8 |
| Column sample by tree | 0.6 | 0.6 | 0.6 |
| Minimum child weight | 5 | 1 | 5 |
| Lambda regularisation | 2 | 10 | 2 |
| Alpha regularisation | 0.1 | 0 | 0.1 |
| Gamma | 0 | 0 | 0 |
| Number of trees | 255 | 27 | 153 |

Of note: regularisation parameters were used in all three models and could mean that covariate reduction has already taken place during feature selection. The CD model, likely due to its small event rate, is the most regularised of the three models. This regularisation was needed to prevent further overfitting.

### Supplementary Note – Description of the inflammatory pathways

The IL-4/IL13 pathway is most enriched through IL-6,[6] TGFB1,[7,8] CCL11,[9,10] IL17A[11,12] and CXCL8.[13] The IL-4/IL-13 pathway has both pro- and anti-inflammatory properties and drives a predominant T-helper 2 (Th2) immune response to pathogens through the recruitment of eosinophils and other leukocytes, and has been implicated in predominantly allergic conditions such as asthma and dermatitis.[7,14]

The proinflammatory IL-23/IL-17 pathway, represented by IL6, IL17A and TGFB1 drives a predominantly neutrophilic immune response through Th17.[11,12,15,16] Th17 mature in response to IL-23, IL-6 and TGF-B to release IL-17A, which plays an important role in neutrophil migration, activation and degranulation, which in turn releases cytokines such as TNF-α, IL-1b and IL-6.

The IL-10 pathway is anti-inflammatory and regulates the IL-4/IL-13 and IL-23/IL-17 pathways via regulatory T-cells (Tregs).[7,17,18] TGF-β drives T-cells to differentiate into Treg cells, which release IL-10 and TGF-β to suppress pro-inflammatory cytokines.[7,18,19] FOXO3 is a transcription factor promoting Treg survival, is associated milder CD,[20] and CSF-1 drives macrophage maturation into anti-inflammatory M2 cells that secrete IL-4, IL-10, and TGF-β.[21,22]

### Supplementary Note – proteins not yet associated with IBD

Of the 48 unique proteins that our model identified, 20 proteins have no prior evidence in existing literature that links them to IBD or its subtypes, CD and UC. Of these 20 proteins, however, 8 proteins have functions which could be plausible mechanisms through which these proteins are associated with IBD, while 12 proteins have no prior evidence of association with IBD, and also have functions which cannot mechanistically explain their association with IBD.

The analysis below provides the reader with context to the proteins' functions and plausible ways in which it can be associated with IBD, but does not seek to claim that these are mechanistically linked to IBD, as further research is needed to prove this association.

These functions are all retrieved from the UniProt website (https://www.uniprot.org) which maintains a large repository of protein functions through gathering existing evidence from mechanistic studies, association studies and other literature.

The 8 proteins that have biological pathways that could explain their association to IBD are: BST, CCL27, TNFRSF14, CST7, COL2A1, SERPINA12, CPPED1 and MZB1.
BST facilitates pre-B-cell growth and is implicated in neutrophil degranulation. CCL27 is a potent chemoattractant of T-lymphocytes in human skin, and IBD is characterised by a predominantly T-cell response. TNFRSF14 is a signalling molecule that promotes immune cell survival and differentiation, and also plays a role in signalling between antigen-presenting cells and lymphocytes. CST7 is implicated in various inflammatory diseases and thought to play a role in immune regulation through inhibition of a unique target in the haematopoietic system. COL2A1 is an ECM protein specific in cartilaginous tissue and ensures normal development of the skeleton. SERPINA12 is an adipokine that modulates insulin action by specifically inhibiting its target protease KLK7 in white adipose tissues. CPPED1 (significantly hazardous in CD) may play an inhibitory role in glucose uptake by adipocytes, blocks cell cycle progression and promotes cell apoptosis. MZB1 acts as a hormone-regulated adipokine/pro-inflammatory cytokine that is implicated in causing chronic inflammation and may have a role in the onset of insulin resistance. MZB1 is thought to play a role in IgA secretion, which is immunosuppressive.

The other 12 proteins whose functions cannot directly be associated with IBD pathogenesis are: F9, BRDT, AMBN, NTF4, CTSV, RGL2, C1QL2, PDIA2, DCTN6, HMCN2, SPINT2 and MSTN.

F9 participates in the intrinsic pathway of blood coagulation by converting factor X to its active form. BRDT is a chromatin protein that binds to histone H4 and is involved in spermatogenesis. AMBN is involved in mineralization and structural organization of enamel. NTF4 is a survival factor for peripheral sensory sympathetic neurons. CTSV is a cysteine protease involved in corneal physiology. RGL2 (significantly protective in CD) is a guanine nucleotide exchange factor that associates with the GTP-bound form of Rap 1A and H-Ras. C1QL2 (significantly protective in CD) may regulate the number of excitatory synapses that are formed on hippocampus neurons. PDIA2 (significantly hazardous in UC) acts as an intracellular oestrogen-binding protein and may act as a chaperone that inhibits aggregation of misfolded proteins. DCTN6 (significantly hazardous in UC) is part of the dynactin complex that activates the molecular motor dynein for ultra-processive transport along microtubules. HMCN2 (significantly protective in UC) has no known functions in human biology yet. SPINT2 inhibits HGF activator which prevents the formation of active hepatocyte growth factor. MSTN is a secreted ligand of the TGF-beta (transforming growth factor-beta) superfamily of proteins, and negatively regulates skeletal muscle cell proliferation and differentiation.

### Supplementary Note – TRIPOD Checklist

This article was reported in accordance with the TRIPOD checklist, which was adapted from the Transparent Reporting of a multivariable prediction model for Individual Prognosis Or Diagnosis (TRIPOD) statement, which is available at https://www.tripod-statement.org.

| Section/Topic | | Checklist Item | Page |
|---|---|---|---|
| Title and abstract | | | |
| Title | 1 | Identify the study as developing and/or validating a multivariable prediction model, the target population, and the outcome to be predicted. | 1 |
| Abstract | 2 | Provide a summary of objectives, study design, setting, participants, sample size, predictors, outcome, statistical analysis, results, and conclusions. | 6 |
| Introduction | | | |
| Background and objectives | 3a | Explain the medical context (including whether diagnostic or prognostic) and rationale for developing or validating the multivariable prediction model, including references to existing models. | 16 - 21 |
| | 3b | Specify the objectives, including whether the study describes the development or validation of the model or both. | 20 - 21 |
| Methods | | | |
| Source of data | 4a | Describe the study design or source of data (e.g., randomized trial, cohort, or registry data), separately for the development and validation data sets, if applicable. | 22 - 23 |
| | 4b | Specify the key study dates, including start of accrual; end of accrual; and, if applicable, end of follow-up. | 22, 24 |
| Participants | 5a | Specify key elements of the study setting (e.g., primary care, secondary care, general population) including number and location of centres. | 22 |
| | 5b | Describe eligibility criteria for participants. | 22 |
| | 5c | Give details of treatments received, if relevant. | N/A |
| Outcome | 6a | Clearly define the outcome that is predicted by the prediction model, including how and when assessed. | 23 -24 |
| | 6b | Report any actions to blind assessment of the outcome to be predicted. | N/A |
| Predictors | 7a | Clearly define all predictors used in developing or validating the multivariable prediction model, including how and when they were measured. | 22, 23, 26, 27 |

| Section/Topic | | Checklist Item | Page |
|---|---|---|---|
| | 7b | Report any actions to blind assessment of predictors for the outcome and other predictors. | N/A |
| Sample size | 8 | Explain how the study size was arrived at. | 22 |
| Missing data | 9 | Describe how missing data were handled (e.g., complete-case analysis, single imputation, multiple imputation) with details of any imputation method. | 28, 30 |
| Statistical analysis methods | 10a | Describe how predictors were handled in the analyses. | 28 - 31 |
| | 10b | Specify type of model, all model-building procedures (including any predictor selection), and method for internal validation. | 27 - 31 |
| | 10d | Specify all measures used to assess model performance and, if relevant, to compare multiple models. | 27 - 31 |
| Risk groups | 11 | Provide details on how risk groups were created, if done. | N/A |
| Results | | | |
| Participants | 13a | Describe the flow of participants through the study, including the number of participants with and without the outcome and, if applicable, a summary of the follow-up time. A diagram may be helpful. | 33 |
| | 13b | Describe the characteristics of the participants (basic demographics, clinical features, available predictors), including the number of participants with missing data for predictors and outcome. | 34 - 35 |
| Model development | 14a | Specify the number of participants and outcome events in each analysis. | 33, 39 |
| | 14b | If done, report the unadjusted association between each candidate predictor and outcome. | N/A |
| Model specification | 15a | Present the full prediction model to allow predictions for individuals (i.e., all regression coefficients, and model intercept or baseline survival at a given time point). | 44,<br>71 - 80 |
| | 15b | Explain how to the use the prediction model. | N/A |
| Model performance | 16 | Report performance measures (with CIs) for the prediction model. | 43 |
| Discussion | | | |
| Limitations | 18 | Discuss any limitations of the study (such as nonrepresentative sample, few events per predictor, missing data). | 51 - 52 |

| Section/Topic | | Checklist Item | Page |
|---|---|---|---|
| Interpretation | 19b | Give an overall interpretation of the results, considering objectives, limitations, and results from similar studies, and other relevant evidence. | 45 - 47 |
| Implications | 20 | Discuss the potential clinical use of the model and implications for future research. | 53 |
| Other information | | | |
| Supplementary information | 21 | Provide information about the availability of supplementary resources, such as study protocol, Web calculator, and data sets. | 3 |
| Funding | 22 | Give the source of funding and the role of the funders for the present study. | 57 |

#### Supplementary Table S1 – Medication codes for NSAID exposure

| Drug name (UKB) | Active NSAID ingredient | UKB coding |
|---|---|---|
| aceclofenac | aceclofenac | 1140925806 |
| preservex 100mg tablet | aceclofenac | 1140925808 |
| acemetacin | acemetacin | 1140875278 |
| emflex 60mg capsule | acemetacin | 1140875282 |
| aspirin | aspirin | 1140868226 |
| aspirin 75mg tablet | aspirin | 1140861806 |
| nu-seals aspirin 75mg e/c tablet | aspirin | 1140864860 |
| caprin 75mg e/c tablet | aspirin | 1140925942 |
| micropirin 75mg e/c tablet | aspirin | 1141177826 |
| solprin 300mg dispersible tablet | aspirin | 1140856214 |
| disprin cv 100mg m/r tablet | aspirin | 1140861808 |
| disprin direct dispersible tablet | aspirin | 1140882192 |
| aspirin+glycine 500mg/133mg dispersible tablet | aspirin | 1140882190 |
| aspirin+codeine 300mg/8mg tablet | aspirin | 1140882268 |
| aspirin+codeine | aspirin | 1140882392 |
| aspirin+methocarbamol 325mg/400mg tablet | aspirin | 1140868282 |
| aspirin+metoclopramide 325mg/5mg effervescent tablet | aspirin | 1140872040 |
| aspirin+papaveretum 500mg/7.71mg dispersible tablet | aspirin | 1141163138 |
| dipyridamole+aspirin | aspirin | 1141167844 |
| isosorbide mononitrate+aspirin | aspirin | 1141164044 |
| azapropazone | azapropazone | 1140871100 |
| rheumox 300mg capsule | azapropazone | 1140871102 |
| celecoxib | celecoxib | 1141176662 |
| celebrex 100mg capsule | celecoxib | 1141176668 |
| celebrex 200mg capsule | celecoxib | 1141176670 |
| dexketoprofen | dexketoprofen | 1141164746 |
| diclofenac | diclofenac | 1140884488 |
| diclofenac sodium+misoprostol | diclofenac | 1140878036 |
| arthrotec tablet | diclofenac | 1140871266 |
| arthrotec 50 tablet | diclofenac | 1140927086 |
| voltarol 25mg e/c tablet | diclofenac | 1140871168 |

| Drug name (UKB) | Active NSAID ingredient | UKB coding |
|---|---|---|
| voltarol 100mg suppository | diclofenac | 1140871174 |
| voltarene 25mg e/c tablet | diclofenac | 1140877868 |
| voltaren retard 100mg m/r tablet | diclofenac | 1140877872 |
| voltarene lp 100mg m/r tablet | diclofenac | 1140877874 |
| volsaid retard 75mg m/r tablet | diclofenac | 1140923920 |
| volraman 25mg e/c tablet | diclofenac | 1140871248 |
| flamrase 25mg e/c tablet | diclofenac | 1140871276 |
| rhumalgan 25mg e/c tablet | diclofenac | 1140871180 |
| econac 100mg suppository | diclofenac | 1141176878 |
| closteril 100 m/r tablet | diclofenac | 1141176886 |
| diflunisal | diflunisal | 1140871282 |
| etodolac | etodolac | 1140871188 |
| etoricoxib | etoricoxib | 1141180140 |
| arcoxia 60mg tablet | etoricoxib | 1141180148 |
| arcoxia 90mg tablet | etoricoxib | 1141180150 |
| arcoxia 120mg tablet | etoricoxib | 1141180152 |
| fenbufen | fenbufen | 1140871202 |
| fenoprofen | fenoprofen | 1140871226 |
| fenopron 300mg tablet | fenoprofen | 1140871228 |
| fenopraine | fenoprofen | 1140910644 |
| flurbiprofen | flurbiprofen | 1140871236 |
| froben 50mg tablet | flurbiprofen | 1140871238 |
| ibuprofen | ibuprofen | 1140871310 |
| ibuprofen product | ibuprofen | 1141157412 |
| ibuprofen+codeine phosphate | ibuprofen | 1140878030 |
| ibuprofen+pseudoephedrine hydrochloride | ibuprofen | 1141184546 |
| propionic acid-ibuprofen | ibuprofen | 1140910496 |
| brufen 200mg tablet | ibuprofen | 1140871374 |
| nurofen 200mg tablet | ibuprofen | 1141187776 |
| anadin ibuprofen 200mg tablet | ibuprofen | 1141153134 |
| advil cold and sinus tablet | ibuprofen | 1141153082 |
| lemsip flu 12hr ibuprofen+pseudoephedrine capsule | ibuprofen | 1141194296 |
| arthrofen 200 tablet | ibuprofen | 1140871320 |
| cuprofen 200mg tablet | ibuprofen | 1140871388 |
| fenbid 300mg spansule | ibuprofen | 1140871394 |
| ibrufhalal 200mg tablet | ibuprofen | 1140877866 |
| indometacin | indometacin | 1140909936 |

| Drug name (UKB) | Active NSAID ingredient | UKB coding |
|---|---|---|
| indomethacin | indometacin | 1140871336 |
| indomethacin product | indometacin | 1141157452 |
| indocid 25mg capsule | indometacin | 1140871354 |
| indolar 25mg capsule | indometacin | 1140853110 |
| imbrilon 25mg capsule | indometacin | 1140871348 |
| ketoprofen | ketoprofen | 1140871506 |
| orudis 50mg capsule | ketoprofen | 1140871516 |
| oruvail 100 m/r capsule | ketoprofen | 1140871522 |
| ketovail 100mg m/r capsule | ketoprofen | 1140871528 |
| ketorolac | ketorolac | 1140884558 |
| mefenamic acid | mefenamic acid | 1140871542 |
| ponstan 250mg capsule | mefenamic acid | 1140871546 |
| meloxicam | meloxicam | 1140926732 |
| mobic 7.5mg tablet | meloxicam | 1140926794 |
| mobic 15mg tablet | meloxicam | 1140926796 |
| nabumetone | nabumetone | 1140875336 |
| relifex 500mg tablet | nabumetone | 1140875338 |
| naproxen | naproxen | 1140871462 |
| naproxen+misoprostol | naproxen | 1140881612 |
| naprosyn 250mg tablet | naproxen | 1140871472 |
| pranoxen continus 375mg m/r tablet | naproxen | 1140871490 |
| napratec tablet combination pack | naproxen | 1140871638 |
| phenylbutazone product | phenylbutazone | 1140871654 |
| parecoxib | parecoxib | 1141180126 |
| piroxicam | piroxicam | 1140871666 |
| piroxicam-beta-cyclodextrin | piroxicam | 1141169526 |
| piroxicam-betadex | piroxicam | 1141182754 |
| feldene 10mg capsule | piroxicam | 1140871672 |
| larapam 10mg capsule | piroxicam | 1140871578 |
| sulindac | sulindac | 1140871604 |
| clinoril 100mg tablet | sulindac | 1140871606 |
| tenoxicam | tenoxicam | 1140875346 |
| mobiflex 20mg tablet | tenoxicam | 1140875350 |
| tiaprofenic acid | tiaprofenic acid | 1140871614 |
| surgam 200mg tablet | tiaprofenic acid | 1140871616 |
| tolmetin | tolmetin | 1140875268 |
| tolectin 200mg capsule | tolmetin | 1140875270 |

#### Supplementary Table S2 – Cox model outputs for all models across the three disease groups: IBD, CD and UC.

##### IBD clinical model:

| Predictor | Hazard ratio (95% CI) | p-value |
|---|---|---|
| Smoker | 1.633 (1.281 – 2.080) | 0.0001 |
| Triglycerides | 1.196 (1.072 – 1.335) | 0.0013 |
| Urban living | 1.082 (0.758 – 1.545) | 0.6630 |
| Age | 1.026 (1.011 – 1.042) | 0.0010 |
| Haemoglobin | 1.026 (0.909 – 1.158) | 0.6810 |
| White cell count | 1.025 (0.983 – 1.069) | 0.2457 |
| CRP | 1.020 (1.003 – 1.038) | 0.0246 |
| Platelet count | 1.003 (1.002 – 1.005) | 0.0002 |
| Vitamin D | 1.003 (0.997 – 1.009) | 0.3572 |
| Creatinine | 1.002 (0.997 – 1.006) | 0.4881 |
| BMI | 0.971 (0.944 – 0.998) | 0.0349 |
| Sex | 0.969 (0.703 – 1.335) | 0.8476 |
| Albumin | 0.962 (0.916 – 1.010) | 0.1160 |
| Cholesterol | 0.844 (0.753 – 0.946) | 0.0035 |
| NSAID exposure | 0.827 (0.631 – 1.085) | 0.1698 |

##### IBD proteomic model:

| Predictor | Hazard ratio (95% CI) | p-value |
|---|---|---|
| PTGDS | 2.719 (1.925 – 3.840) | 0.0000 |
| ORM1 | 1.910 (1.004 – 3.633) | 0.0486 |
| F9 | 1.569 (0.818 – 3.009) | 0.1748 |
| CCL11 | 1.547 (1.146 – 2.088) | 0.0044 |
| MMP12 | 1.248 (1.032 – 1.510) | 0.0225 |
| TGFA | 1.169 (0.925 – 1.477) | 0.1912 |
| CXCL9 | 1.142 (0.986 – 1.323) | 0.0757 |
| IL6 | 1.142 (1.007 – 1.294) | 0.0390 |
| CCL27 | 1.141 (0.945 – 1.376) | 0.1702 |
| COL2A1 | 1.129 (0.952 – 1.338) | 0.1636 |
| IL17A | 1.099 (0.881 – 1.372) | 0.4025 |
| TGFB1 | 1.067 (0.744 – 1.530) | 0.7249 |
| SEL1L | 1.043 (0.686 – 1.585) | 0.8434 |
| CXCL8 | 1.033 (0.871 – 1.225) | 0.7088 |
| NECTIN2 | 1.009 (0.630 – 1.615) | 0.9704 |
| ASGR1 | 0.883 (0.576 – 1.353) | 0.5672 |

| Predictor | Hazard ratio (95% CI) | p-value |
|---|---|---|
| APOBR | 0.714 (0.540 – 0.946) | 0.0188 |
| TNXB | 0.694 (0.456 – 1.057) | 0.0891 |
| DPP4 | 0.483 (0.331 – 0.704) | 0.0002 |
| COL15A1 | 0.318 (0.216 – 0.469) | 0.0000 |

#### IBD combined model:

| Predictor | Hazard ratio (95% CI) | p-value |
|---|---|---|
| PTGDS | 3.002 (2.100 – 4.293) | 0.0000 |
| ORM1 | 1.973 (0.984 – 3.955) | 0.0556 |
| F9 | 1.739 (0.880 – 3.435) | 0.1111 |
| CCL11 | 1.485 (1.086 – 2.030) | 0.0132 |
| Smoker | 1.477 (1.151 – 1.894) | 0.0021 |
| TGFA | 1.261 (0.965 – 1.649) | 0.0896 |
| MMP12 | 1.228 (1.006 – 1.499) | 0.0434 |
| IL6 | 1.192 (1.053 – 1.349) | 0.0054 |
| CCL27 | 1.134 (0.921 – 1.396) | 0.2371 |
| CXCL9 | 1.125 (0.968 – 1.308) | 0.1241 |
| SEL1L | 1.116 (0.745 – 1.673) | 0.5945 |
| COL2A1 | 1.115 (0.940 – 1.323) | 0.2109 |
| IL17A | 1.112 (0.890 – 1.388) | 0.3512 |
| Haemoglobin | 1.064 (0.945 – 1.197) | 0.3058 |
| TGFB1 | 1.035 (0.713 – 1.501) | 0.8579 |
| Urban living | 1.029 (0.720 – 1.470) | 0.8763 |
| CXCL8 | 1.029 (0.863 – 1.227) | 0.7524 |
| Triglycerides | 1.026 (0.900 – 1.170) | 0.7027 |
| Albumin | 1.008 (0.959 – 1.058) | 0.7639 |
| Vitamin D | 1.004 (0.999 – 1.010) | 0.1279 |
| Platelet count | 1.001 (0.999 – 1.003) | 0.1550 |
| NECTIN2 | 0.998 (0.608 – 1.640) | 0.9941 |
| Age | 0.996 (0.979 – 1.013) | 0.6265 |
| Creatinine | 0.995 (0.987 – 1.003) | 0.2621 |
| CRP | 0.984 (0.960 – 1.009) | 0.2015 |
| BMI | 0.982 (0.953 – 1.012) | 0.2362 |
| ASGR1 | 0.971 (0.619 – 1.522) | 0.8971 |
| White cell count | 0.946 (0.875 – 1.023) | 0.1656 |
| Cholesterol | 0.925 (0.823 – 1.039) | 0.1886 |
| Sex | 0.841 (0.602 – 1.176) | 0.3109 |
| NSAID exposure | 0.779 (0.593 – 1.024) | 0.0738 |

| Predictor | Hazard ratio (95% CI) | p-value |
|---|---|---|
| APOBR | 0.709 (0.535 – 0.940) | 0.0167 |
| TNXB | 0.653 (0.423 – 1.008) | 0.0541 |
| DPP4 | 0.458 (0.308 – 0.680) | 0.0001 |
| COL15A1 | 0.388 (0.241 – 0.623) | 0.0001 |

#### CD clinical model:

| Predictor | Hazard ratio (95% CI) | p-value |
|---|---|---|
| Smoker | 1.358 (0.895 – 2.062) | 0.1500 |
| Triglycerides | 1.241 (1.025 – 1.502) | 0.0270 |
| Urban living | 1.116 (0.593 – 2.101) | 0.7338 |
| Age | 1.039 (1.011 – 1.067) | 0.0067 |
| CRP | 1.021 (0.994 – 1.047) | 0.1264 |
| Platelet count | 1.004 (1.001 – 1.007) | 0.0209 |
| Creatinine | 1.003 (0.999 – 1.008) | 0.1158 |
| Vitamin D | 1.000 (0.990 – 1.010) | 0.9948 |
| White cell count | 0.993 (0.890 – 1.109) | 0.9036 |
| BMI | 0.969 (0.925 – 1.014) | 0.1728 |
| Haemoglobin | 0.960 (0.783 – 1.177) | 0.6922 |
| Sex | 0.900 (0.527 – 1.539) | 0.7015 |
| Albumin | 0.898 (0.828 – 0.974) | 0.0091 |
| NSAID exposure | 0.889 (0.563 – 1.405) | 0.6150 |
| Cholesterol | 0.738 (0.603 – 0.904) | 0.0033 |

#### CD proteomic model:

| Predictor | Hazard ratio (95% CI) | p-value |
|---|---|---|
| CSF1 | 1.734 (0.733 – 4.101) | 0.2104 |
| SPINT2 | 1.485 (0.786 – 2.806) | 0.2235 |
| REG1B | 1.389 (1.019 – 1.893) | 0.0374 |
| BST1 | 1.359 (0.829 – 2.229) | 0.2243 |
| CPPED1 | 1.339 (1.005 – 1.784) | 0.0461 |
| DEFA1_DEFA1B | 1.295 (0.977 – 1.716) | 0.0722 |
| IL6 | 1.273 (1.051 – 1.542) | 0.0136 |
| REG3A | 1.196 (0.868 – 1.648) | 0.2728 |
| TNFRSF14 | 1.049 (0.423 – 2.601) | 0.9181 |
| FOXO3 | 1.028 (0.822 – 1.285) | 0.8120 |
| CXCL8 | 1.005 (0.758 – 1.332) | 0.9748 |
| LBP | 0.968 (0.703 – 1.333) | 0.8411 |
| DLL1 | 0.768 (0.345 – 1.711) | 0.5178 |

| Predictor | Hazard ratio (95% CI) | p-value |
|---|---|---|
| NTF4 | 0.720 (0.386 – 1.346) | 0.3038 |
| MSTN | 0.704 (0.508 – 0.976) | 0.0352 |
| CTSV | 0.654 (0.413 – 1.036) | 0.0707 |
| ITGA11 | 0.634 (0.377 – 1.066) | 0.0855 |
| RGL2 | 0.599 (0.360 – 0.997) | 0.0486 |
| APOBR | 0.574 (0.358 – 0.920) | 0.0210 |
| C1QL2 | 0.466 (0.273 – 0.797) | 0.0053 |

#### CD combined model:

| Predictor | Hazard ratio (95% CI) | p-value |
|---|---|---|
| CSF1 | 1.886 (0.757 – 4.697) | 0.1729 |
| SPINT2 | 1.476 (0.768 – 2.836) | 0.2427 |
| REG1B | 1.449 (1.055 – 1.990) | 0.0221 |
| DEFA1_DEFA1B | 1.439 (1.089 – 1.900) | 0.0103 |
| TNFRSF14 | 1.382 (0.498 – 3.838) | 0.5343 |
| BST1 | 1.373 (0.826 – 2.282) | 0.2212 |
| CPPED1 | 1.351 (1.007 – 1.813) | 0.0445 |
| IL6 | 1.318 (1.095 – 1.586) | 0.0035 |
| Smoker | 1.313 (0.861 – 2.004) | 0.2064 |
| REG3A | 1.133 (0.813 – 1.580) | 0.4612 |
| Triglycerides | 1.122 (0.909 – 1.383) | 0.2836 |
| LBP | 1.069 (0.759 – 1.505) | 0.7035 |
| Urban living | 1.021 (0.540 – 1.927) | 0.9500 |
| Age | 1.007 (0.978 – 1.037) | 0.6381 |
| Vitamin D | 1.004 (0.994 – 1.014) | 0.4359 |
| Platelet count | 1.001 (0.998 – 1.004) | 0.5492 |
| FOXO3 | 0.999 (0.792 – 1.260) | 0.9929 |
| Creatinine | 0.993 (0.983 – 1.003) | 0.1565 |
| Haemoglobin | 0.984 (0.803 – 1.205) | 0.8743 |
| CRP | 0.975 (0.939 – 1.012) | 0.1795 |
| Albumin | 0.961 (0.886 – 1.042) | 0.3338 |
| BMI | 0.955 (0.912 – 1.001) | 0.0543 |
| CXCL8 | 0.920 (0.676 – 1.252) | 0.5960 |
| White cell count | 0.908 (0.805 – 1.025) | 0.1178 |
| NSAID exposure | 0.902 (0.570 – 1.428) | 0.6601 |
| Sex | 0.885 (0.481 – 1.628) | 0.6941 |
| Cholesterol | 0.880 (0.719 – 1.078) | 0.2165 |
| MSTN | 0.808 (0.554 – 1.179) | 0.2697 |

| Predictor | Hazard ratio (95% CI) | p-value |
|---|---|---|
| NTF4 | 0.748 (0.403 – 1.389) | 0.3579 |
| DLL1 | 0.700 (0.299 – 1.638) | 0.4103 |
| CTSV | 0.657 (0.403 – 1.072) | 0.0924 |
| APOBR | 0.581 (0.362 – 0.934) | 0.0248 |
| RGL2 | 0.578 (0.344 – 0.972) | 0.0385 |
| ITGA11 | 0.552 (0.317 – 0.962) | 0.0361 |
| C1QL2 | 0.451 (0.256 – 0.793) | 0.0057 |

#### UC clinical model:

| Predictor | Hazard ratio (95% CI) | p-value |
|---|---|---|
| Smoker | 1.791 (1.328 – 2.414) | 0.0001 |
| Triglycerides | 1.175 (1.028 – 1.342) | 0.0178 |
| Haemoglobin | 1.069 (0.919 – 1.243) | 0.3873 |
| Urban living | 1.064 (0.692 – 1.637) | 0.7770 |
| Sex | 1.056 (0.698 – 1.599) | 0.7959 |
| White cell count | 1.033 (0.992 – 1.076) | 0.1173 |
| Age | 1.021 (1.002 – 1.041) | 0.0269 |
| CRP | 1.019 (0.995 – 1.043) | 0.1205 |
| Vitamin D | 1.004 (0.997 – 1.011) | 0.2264 |
| Platelet count | 1.003 (1.001 – 1.006) | 0.0038 |
| Albumin | 0.997 (0.939 – 1.059) | 0.9240 |
| Creatinine | 0.997 (0.986 – 1.007) | 0.5032 |
| BMI | 0.974 (0.941 – 1.008) | 0.1342 |
| Cholesterol | 0.897 (0.781 – 1.029) | 0.1218 |
| NSAID exposure | 0.797 (0.569 – 1.116) | 0.1862 |

#### UC proteomic model:

| Predictor | Hazard ratio (95% CI) | p-value |
|---|---|---|
| PTGDS | 2.715 (1.903 – 3.874) | 0.0000 |
| PDIA2 | 1.662 (1.300 – 2.123) | 0.0000 |
| MMP12 | 1.414 (1.116 – 1.791) | 0.0041 |
| CCL11 | 1.388 (0.964 – 1.998) | 0.0778 |
| COL2A1 | 1.227 (0.961 – 1.567) | 0.1011 |
| TGFA | 1.219 (0.925 – 1.608) | 0.1597 |
| DCTN6 | 1.197 (1.019 – 1.407) | 0.0288 |
| AGR2 | 1.121 (0.984 – 1.278) | 0.0858 |
| CST7 | 1.113 (0.981 – 1.261) | 0.0957 |

| Predictor | Hazard ratio (95% CI) | p-value |
|---|---|---|
| CXCL9 | 1.053 (0.876 – 1.266) | 0.5815 |
| MZB1 | 1.046 (0.812 – 1.347) | 0.7284 |
| SERPINA12 | 0.975 (0.866 – 1.097) | 0.6761 |
| PDGFA | 0.968 (0.737 – 1.270) | 0.8123 |
| SEL1L | 0.924 (0.556 – 1.537) | 0.7618 |
| CA6 | 0.874 (0.718 – 1.064) | 0.1799 |
| HMCN2 | 0.730 (0.570 – 0.936) | 0.0130 |
| BRDT | 0.685 (0.447 – 1.051) | 0.0829 |
| TNXB | 0.668 (0.404 – 1.106) | 0.1172 |
| AMBN | 0.621 (0.353 – 1.095) | 0.0997 |
| COL15A1 | 0.346 (0.218 – 0.548) | 0.0000 |

#### UC combined model:

| Predictor | Hazard ratio (95% CI) | p-value |
|---|---|---|
| PTGDS | 2.994 (2.088 – 4.295) | 0.0000 |
| PDIA2 | 1.687 (1.306 – 2.179) | 0.0001 |
| Smoker | 1.605 (1.181 – 2.180) | 0.0025 |
| MMP12 | 1.361 (1.067 – 1.737) | 0.0132 |
| CCL11 | 1.317 (0.899 – 1.929) | 0.1573 |
| TGFA | 1.259 (0.918 – 1.727) | 0.1534 |
| COL2A1 | 1.232 (0.961 – 1.578) | 0.0999 |
| Sex | 1.182 (0.763 – 1.829) | 0.4543 |
| DCTN6 | 1.181 (1.000 – 1.393) | 0.0497 |
| AGR2 | 1.144 (1.002 – 1.305) | 0.0472 |
| CST7 | 1.102 (0.970 – 1.252) | 0.1355 |
| CXCL9 | 1.089 (0.904 – 1.311) | 0.3689 |
| SEL1L | 1.064 (0.656 – 1.727) | 0.8005 |
| Haemoglobin | 1.050 (0.908 – 1.213) | 0.5099 |
| Triglycerides | 1.048 (0.906 – 1.212) | 0.5286 |
| Urban living | 1.048 (0.681 – 1.613) | 0.8317 |
| MZB1 | 1.048 (0.800 – 1.372) | 0.7351 |
| Albumin | 1.040 (0.980 – 1.104) | 0.1919 |
| Vitamin D | 1.006 (0.999 – 1.013) | 0.0981 |
| Platelet count | 1.002 (0.999 – 1.004) | 0.2122 |
| CRP | 0.999 (0.971 – 1.028) | 0.9505 |
| SERPINA12 | 0.997 (0.886 – 1.123) | 0.9618 |
| BMI | 0.995 (0.961 – 1.031) | 0.7908 |
| Age | 0.992 (0.972 – 1.012) | 0.4419 |

| Predictor | Hazard ratio (95% CI) | p-value |
|---|---|---|
| Creatinine | 0.991 (0.979 – 1.002) | 0.1181 |
| White cell count | 0.986 (0.910 – 1.067) | 0.7227 |
| Cholesterol | 0.971 (0.844 – 1.116) | 0.6754 |
| PDGFA | 0.920 (0.695 – 1.217) | 0.5574 |
| CA6 | 0.845 (0.682 – 1.047) | 0.1234 |
| HMCN2 | 0.745 (0.579 – 0.959) | 0.0223 |
| NSAID exposure | 0.741 (0.527 – 1.042) | 0.0852 |
| BRDT | 0.654 (0.424 – 1.009) | 0.0547 |
| TNXB | 0.621 (0.368 – 1.047) | 0.0737 |
| AMBN | 0.618 (0.348 – 1.099) | 0.1014 |
| COL15A1 | 0.423 (0.237 – 0.754) | 0.0036 |

### Supplementary Table S3 – Proteins shared between the proteomics models and existing models

**Supplementary Table S3 – Proteins shared between our proteomics models and existing models.** The IBD model shared the most proteins. In contrast, the UC and CD models shared far fewer proteins.

**AGR2**, anterior gradient 2; **APOBR**, apolipoprotein B receptor; **CCL11**, C-C motif chemokine ligand 11; **COL15A1**, collagen type XV alpha 1 chain; **CSF1**, colony-stimulating factor 1; **CXCL9**, C-X-C motif chemokine ligand 9; **DEFA1**, defensin alpha 1; **IL6**, interleukin-6; **ITGA11**, integrin subunit alpha 11; **MMP12,** matrix metalloproteinase-12; **REG1B**, regenerating islet-derived protein 1-beta.

| **Disease group** | **Study (authors)** | **Shared proteins with our model** | **Number of shared proteins** |
|---|---|---|---|
| IBD | Zhang et. al.[23] | MMP12, CCL11, COL15A1, AGR2, APOBR, CXCL9, IL6 | 7 out of 20 |
| UC | Zhang et. al.[23] | MMP12, CCL11 | 2 out of 20 |
| CD | Zhang et. al.[23] | CSF1, REG1B | 2 out of 20 |
| | Feng et. al.[24] | REG1B, ITGA11, DEFA1 | 3 out of 20 |
